# Statistical Analysis Plan

**DOI:** 10.64898/2026.03.11.26348100

**Authors:** Julie Bernhardt, Leonid Churilov, the AVERT-DOSE Collaboration

**Affiliations:** The University of Melbourne; Florey Institute of Neuroscience and Mental Health

## Abstract

This paper reports the statistical analysis plan for the AVERT-DOS trial: A Phase 3, Multi Arm, Multi Stage, Covariate Adjusted, Response Adaptive, Randomised Trial to Determine Optimal Early Mobility Training after Stroke. It contains a trial overview, regulatory information and details of the planned main analyses for the study protocol version Version 5; 15 June 2025.

## 3. Administrative Information

Main Protocol: 001-1; Version 5; 15 June 2025

Interventional Protocol: 001-2 Version 4 1 July 2024

ANZCTR register Identifier: ACTRN12619000557134

### 3.1 Document Version History

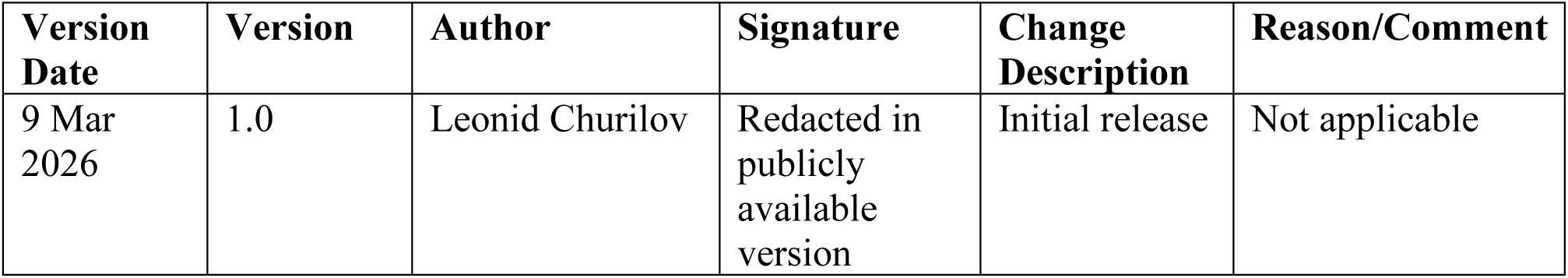

### 3.2 Approvals

The undersigned have reviewed this plan and approve it as final. They find it to be consistent with the requirements of the protocol as it applies to their respective areas. They also find it to be compliant with ICH-E9 principles and confirm that this analysis plan was developed in a completely blinded manner (i.e., without knowledge of the effect of the intervention being assessed)

| Name | Role on Study | Affiliation | Signature | Date |
| --- | --- | --- | --- | --- |
| Julie Bernhardt | PI | Florey Institute of Neuroscience and Mental Health Austin Campus | Redacted in publicly available version | 12 Mar 2026 |
| Leonid Churilov | Biostatistics Lead | University of Melbourne | Redacted in publicly available version | 12 Mar 2026 |

## 4. Study design

### 4.1 Study overview

AVERT-DOSE was designed as an international, multi-centre, dose-finding, Multi-Arm (three intervention arms and a pre-specified reference arm), Multi-Stage (2 stages), Covariate-Adjusted, Response-Adaptive (MAMS CARA) randomised trial with blinded outcome assessments at 3 and 6 months post stroke. The trial aims to identify the best early mobility based intervention regimen for individuals with both mild and moderate stroke severity, considered as two separate strata. The original MAMS CARA design is outlined below and illustrated in Figure 1a. The design allows to test multiple promising intervention regimens at trial start, with the treatment effects (favourable outcome, mRS 0-2) of each intervention regimen dynamically informing the probability of allocation of participants to subsequent interventions as the trial progresses in an automated and pre-specified manner. Within each stratum, at the end of Stage 1, the single best-performing intervention arm together with the pre-specified reference arm were to be taken to Stage 2 to identify the optimal intervention regimen. The original sample size was planned as 2,700 participants (1300 mild stratum; 1400 moderate stratum) with potential adaptive sample size re-estimation for up to the maximum of 3,600 participants (1600 mild stratum; 1900 moderate stratum).

**Figure 1a.**
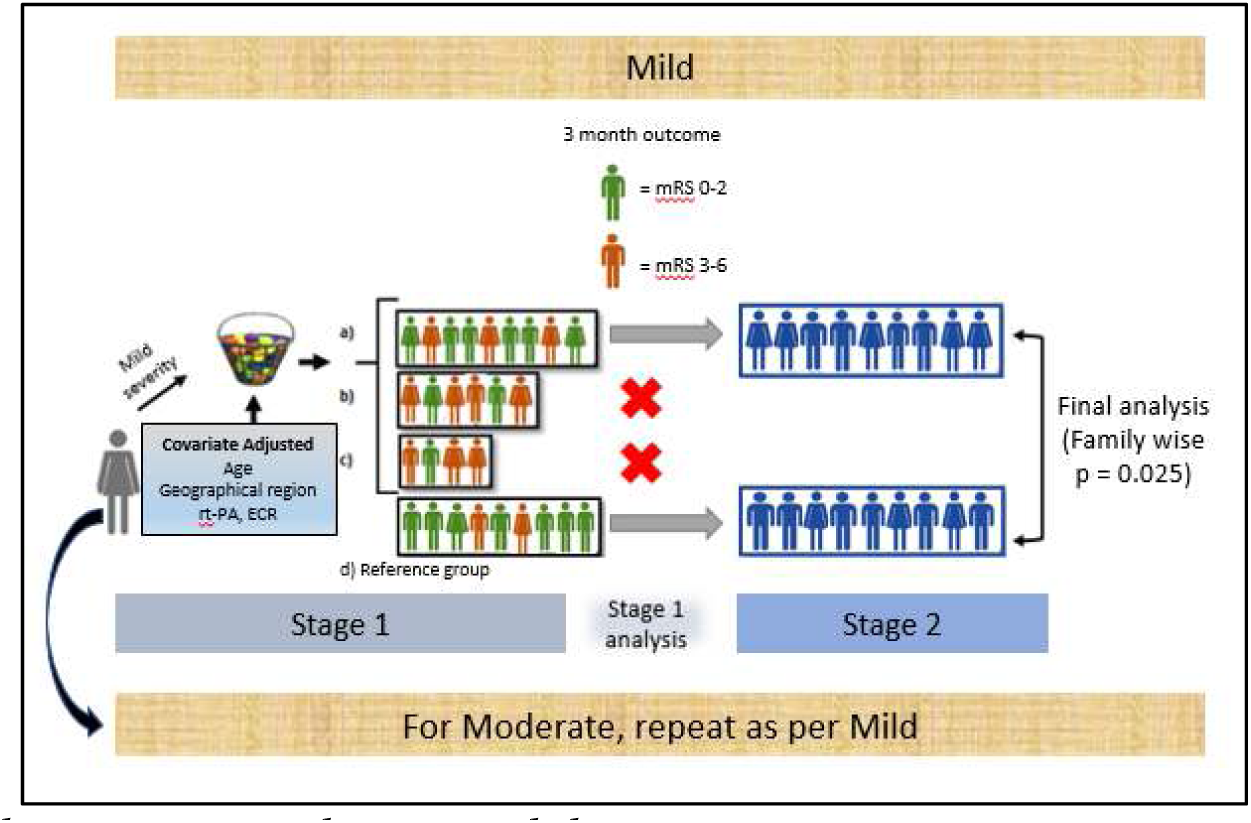
Full schematic as per the original design.

The experimental intervention is task-specific, upright or out of bed sitting, standing and walking training varying in amount, frequency, task difficulty and intensity starting within 48 hours of stroke onset and continued for 14 days or until discharge from acute stroke care (whichever is sooner). The task-specific training is delivered by trained nurses/physiotherapists. Post stroke functional ability according to stroke severity strata (NIHSS) dictates intervention dose with 4 titrations specified for each stratum, which includes a low dose reference arm. There is no ‘usual care’ control in this study. Frequency and minutes of intervention are controlled in each treatment arm, while activity type and intensity of training (i.e., how hard the patient is working during the intervention) is adjusted in line with recovery as dictated by the therapist. Nursing staff supported functional training at specified doses according to protocol. All interventions are recorded online. Intervention staff received feedback on dose compliance. Patients are followed up at 3 and 6 months in a blinded manner.

In early 2025 it became apparent that the trial would be stopped early due to funding constraints. The design was modified in a blinded manner from a four-arm two-stage CARA (figure 1a) to a four-arm single stage CARA. This is reflected in the most recent version of the study Protocol (001-1, Version 5.0, Date: 15 June 2025) and shown in Figure 1b. The final sample size was planned as approximately 1000 participants (650 mild stratum; 350 moderate stratum).

**Figure 1b.**
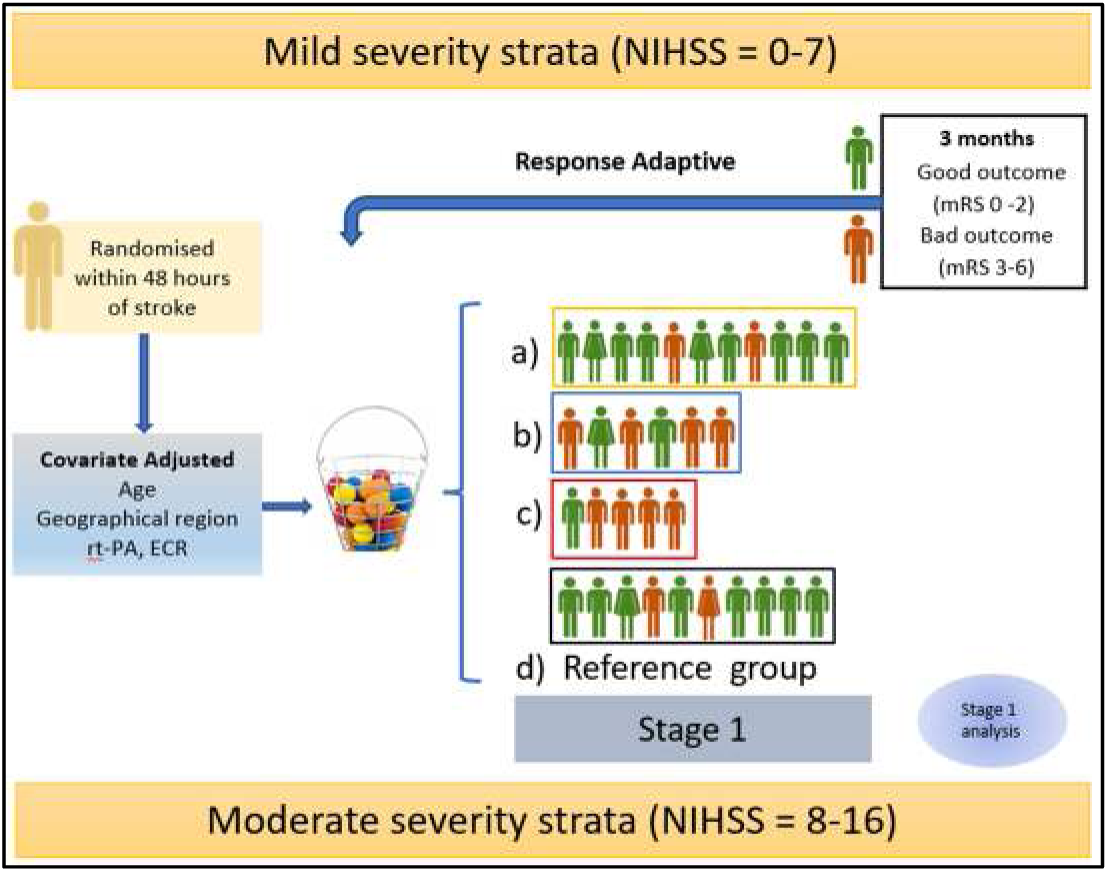
In this figure good outcome on the mRS is shown in Stage 1 with green figures, poor outcomes with red figures. In this example, treatment (a) is more beneficial in outcome (more participants achieve mRS 0-2; no or little disability at 3 months) and so the number of participants allocated to this group is increased. This process occurs iteratively as new outcomes become known.

### 4.2 Aims and hypotheses

#### 4.2.1 Primary

The main aim of this study is to define the optimal early intervention regimens for people with ischaemic stroke of mild and moderate severity. We hypothesise that against a pre-specified reference group, the optimal dose intervention regimen(s) will result in more participants experiencing favourable outcome (mRS 0-2) at 3 months.

#### 4.2.2 Secondary

We **hypothesise** that against a pre-specified reference group, the optimal dose intervention regimen(s) will result in:

- Fewer deaths at 14 days post randomisation and at 3 months
- Participants experiencing fewer and less severe complications during the intervention period (< 14 days post randomisation)
- Faster time to unassisted walking 50 metres within 3 months
- Increased walking speed at 3 months
- Better quality of life at 3 months *and*
- More cost-effective care at 6 months

#### 4.2.3 Sub studies

The pre-planned studies within a trial (sub studies) were to include:

- Genome Wide Association analysis and stroke recovery;
- Long term stroke service usage using data linkage; and
- Prior cognitive reserve and favourable outcome.

The analytical plans for these studies are not included in this SAP document and will be presented in separate relevant documents.

### 4.3 Patient population

Participants were recruited from various geographic regions including Australasia, UK/Europe, South America, Asia (India and South East Asia). Hospitals with a geographically co-located stroke unit were selected for participation in this study.

#### Inclusion criteria

Patients admitted to stroke unit with:

- Ischaemic stroke (first ever or recurrent)
- Aged ≥ 18 years
- Ability to be enrolled within 48 hours of the onset of stroke symptoms. (Stroke onset is defined as the date and time the patient was known to be symptom free)
- Mild (NIHSS 0-7) or moderate stroke severity (NIHSS 8 ≤ 16)
- Pre stroke mRS of 0 – 2
- Participants are medically stable at time of recruitment (i.e. meet physiological criteria: (participant rousable, SBP > 120 mmHg and < 180 mmHg, O2 saturation > 92%, HR > 40 and < 100, and temperature < 38.5°C).

##### Exclusion Criteria

Patients with:

- Pre-stroke mRS of 3, 4 or 5 (indicating moderate to severe pre-morbid disability)
- Diagnosis of haemorrhagic stroke or transient ischaemic attack
- Severe stroke (NIHSS > 16)
- Co-morbid progressive neurological conditions
- Severe heart failure, unstable coronary, or any other condition that is judged by the investigator to impose a hazard to the participant if involved in the trial (including COVID 19)
- Concurrent diagnosis of rapidly deteriorating disease (e.g. terminal cancer)
- Deterioration following admission, resulting in a documented clinical decision for palliative treatment, or immediate surgery
- A lower limb fracture or other disability which deems the participant unable to participate in mobility training
- Patients with no evident mobility problems
- Patients expected to be discharged within 3 days of trial enrolment
- Current participation in a drug or other intervention trial

##### Additional inclusion notes

- Participants may receive thrombolytic and/or ECR by 48 hours post stroke
- Participants may be included in observational trial/s (i.e. no intervention provided) with approval by the trial sponsor.
- Participants can be included if they are admitted via intensive care or other hyper acute unit as long as there are plans for transfer to the acute stroke unit within 48 hours of stroke.
- Time of onset for wake up strokes will be taken from when the patient was last seen well.

### 4.4 Randomisation

#### 4.4.1 Overview

Patients are randomised in a covariate-adjusted response adaptive (CARA) manner. Randomisation is undertaken independently for each patient and is stratified by stroke severity. Mild stroke is defined as a baseline NIHSS score of 1-7, and moderate stroke is defined as a baseline NIHSS score of 8-16. Data from one stratum are not used by the adaptive randomisation procedure within the other stratum.

#### 4.4.2 Technical Details

The CARA randomisation procedure consists of a covariate-adjusted and response-adaptive component.

##### Covariate-Adjusted Randomisation Component of the Design

The covariate adjusted component of the randomisation procedure balances age (defined in bins of 18-64; 65-80; and 81+), region (Aus; Asia; UK/Europe; and South America) and hyperacute treatment (neither thrombolysis nor endovascular clot retrieval; Thrombolysis but not endovascular clot retrieval; endovascular clot retrieval but not thrombolysis; and both thrombolysis and endovascular clot retrieval) across the four treatment arms. This component of the randomisation procedure produces a probability of treatment assignment using Pocock and Simon^(1)^ biased coin minimisation.

For each covariate and each treatment arm, the covariate-adjusted component of the randomisation procedure calculates the prospective imbalance in treatment assignment that would occur if that covariate were a participant to be randomised to each arm. Imbalance is measured by first calculating the number of participants in each group that match a given covariate (e.g. if the new participant is aged 18-64, the count of people aged 18-64 across the four arms is used), then calculating the standard deviation of these counts. The total prospective imbalance for each arm is calculated by summing the standard deviation of these counts across the three covariates.

The probability of assignment to each arm is then given using the formula:

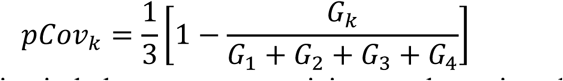

where *G_k_* is the total prospective imbalance were a participant to be assigned to arm *k*. If the total prospective imbalance in one arm is lower than the others, the probability of treatment assignment to that arm is higher.

##### Response-Adaptive Randomisation Component

The response adaptive component of the randomisation procedure sends additional participants to arms that have more recorded positive outcomes (defined as mRS 0-2 at 90 days) and fewer participants to arms that have more recorded negative outcomes (defined as mRS 6 at 90 days). It uses an *Urn Model*^(2)^ to achieve this. An *Urn Model* conceptualises randomisation as the process of randomly selecting with replacement marked balls from an urn (with marking indicating arm). The probability of treatment assignment is modified by changing the number of balls of each type in the urn. Each arm begins with 100 balls. For every positive outcome observed in that arm, one ball is added. For every negative outcome, one ball is removed. The probability of assignment to each arm based on the urn model is therefore given by:

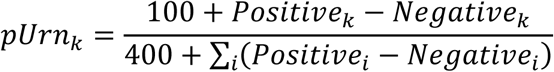

where *Positive_k_* is the count of participants in arm *k* with mRS 0-2 and *Negative_k_* is the count of participants in arm *k* with mRS 6. This probability is then modified to fix the probability of the reference arm to be 25%.

The final response-adaptive randomisation probability is given by:

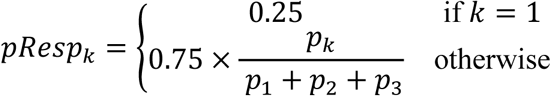

where *k*=1 represents the reference arm.

##### Covariate-Adjusted Response Adaptive Randomisation

The final probability of treatment assignment is given by multiplying these probabilities together and rescaling to sum to 1, resulting in:

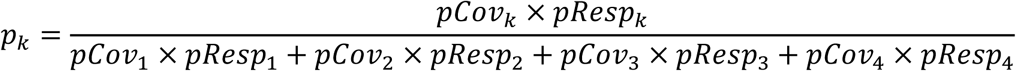

This corresponds to the approach taken by Ning & Huang^(3)^ generalised to more than two groups.

##### Implementation

This randomisation algorithm was implemented in PHP and interfaces with the AVERT-DOSE REDCap eCRF. All MAMS CARA codes and settings were fully prespecified, written and implemented prior to the start of the trial. An independent unblinded statistician, different to the study statistician, monitored the performance of the randomisation process. As the trial progressed, trial staff were prevented from identifying reference or optimal groups as adaptive randomisation occurred across sites. The Data Safety and Monitoring Committee were unblinded.

### 4.5 Baseline and follow-up assessments

All responsible investigators receive training in the systems for data collection and data entry and training in Good Clinical Practice (GCP). All investigators responsible for patient assessment participate in credentialled training for critical assessment items (FMA, NIHSS, mRS).

The assessment schedule for this trial is outlined in the main study protocol (001-1 V5 15 June 2025).

Participants undergo a series of baseline assessments, during the acute stroke admission and receive allocated intervention during this time for up to 14 days. Intervention is outlined in the study intervention protocol (001-2 Version 4, 1 July 2024), provided only to investigators responsible for the delivery of protocolised therapy with participants. Participants are randomised to receive one of four doses of out of bed training per severity strata (mild NIHSS 0 – 7, Moderate NIHSS 8 – 16), with one dose the pre-specified reference arm.

Adverse events are collected during the intervention phase, important medical events collected until 3 months, and Serious Adverse Events collected until 6 months. The 3 and 6 month assessments of post-stroke disability (mRS) and secondary outcome measures are conducted by trained staff blinded to group allocation and the therapy protocol.

### 4.6 Sample size considerations

As discussed in detail in the Definitions of the Primary Estimand and Outcomes (section 3.8), the primary outcome for this study is the proportion of participants achieving favourable outcome (independence, no or little disability; modified Rankin Scale (mRS) score 0-2).

The sample size estimation for the original four-arm two-stage design was undertaken using the probabilities of favourable outcome observed in AVERT trial data^(4)^ for the different doses proposed in this study, extensive simulations have been run for Mild and Moderate stroke severity strata to identify the sample sizes to yield 80% power to observe 10% absolute treatment effects or larger compared to a pre-specified reference group in both severity strata (assuming 70% incidence of favourable outcome in a pre-specified reference group in both severity strata assuming Bonferroni corrected one-sided alpha family-wise threshold p=0.025 per stratum). Recruiting 1300 participants in Mild stratum and 1400 participants in Moderate stratum would yield 80% power.

Adaptive sample size re-estimation were to be undertaken for individual strata using Mehta and Pocock promising zone methodology^(5)^ with a potential increase of up to the maximum of 1600 participants for Mild stroke stratum and 1900 participants for Moderate stroke stratum.

While original sample size estimate forecast recruitment to 2,700 participants, with potential adaptive sample size re-estimation half-way through the trial, the COVID-19 pandemic led to a major disruption to study recruitment. Current recruitment numbers and termination of funding (31 Dec 2025) meant we had insufficient numbers of participants to execute the pre-planned adaptive sample size re-estimation and/or achieve the pre-planned sample size. In early 2025 it became apparent that the trial would be stopped early. The design was modified from a four-arm two-stage CARA to a four-arm single stage CARA in a blinded manner as reflected in the most recent version of the study Protocol (No: 001-1 Version 5.0, Date: 15 June 2025) and shown in Figure 1b. The final sample size was planned as approximately 1,000 participants (650 mild stratum; 350 moderate stratum).

The last participant was recruited on 4 December 2025, resulting in the total achieved sample size of 1000 participants.

### 4.7 Unblinding

Participants were allocated to one of the four groups of mobility training without the knowledge of what the reference group comprises. Participants and family/carers were not told of the group allocation. Trial physiotherapists and nurses were aware of their group allocation as they are delivering the interventions but they were blinded to which group comprises the reference group and which is the predicted optimal group. Other ward staff were not told of group allocation so as to not bias usual processes of care. Trial staff did not record the daily dose schedule on the participant’s medical record. Blinding of the outcomes assessor was maintained by using off-site trained assessors to collect all outcome data. Treatment allocations were securely stored and separated from the assessors involved in all assessments. Statisticians not involved in the DSMB remain blinded and work on dummy datasets until the computer codes for statistical analysis are validated. Only the Data and Safety Monitoring Board (DSMB) have access to interim data and results. The DSMB reviewed unblinded data in accordance with the DSMB Charter.

### 4.8 Definitions of Primary Estimand and Secondary and Exploratory Outcomes

This trial follows the Estimand framework for the design and analysis of clinical trials. The objective of the Estimand framework as presented in addendum (R1) to the International Council on Harmonization E9 guidance^(6)^ is to align the clinical trial objectives with the study design, endpoints and analysis, in order to improve study planning and the interpretation of the study results.

The Estimand framework requires appropriate pre-specification of population, individual-level outcome measure, population level summary measure, and strategies for handling intercurrent events^(7)^ The trial Primary Estimand and secondary and exploratory outcomes are described below while respective analysis methods are detailed in Statistical Analysis section of this SAP.

#### 4.8.1 Primary Estimand: achieving little or no disability

**Population:** All included participants as reflected by the trial inclusion and exclusion criteria who did not withdraw consent prior to treatment initiation.

**Individual-level outcome measure:** achieving a favourable outcome of no or little disability (mRS score 0-2) at 3 months post stroke. The mRS enables categorisation of participants based on 7 levels (0-6), ordered for dependency and the ability to look after themselves in daily life, with death included as an outcome. It is used as the primary outcome in large stroke clinical trials,^(8)^ and is recommended as a core measure in rehabilitation and recovery trials.

**Population-level summary outcome measure:** the proportion of participants achieving a favourable outcome of no or little disability.

**Strategy for Intercurrent events:** *Treatment policy strategy* – any intercurrent events which prevent collection of the primary outcome measure will be treated according to a treatment policy strategy. Missingness mechanism will be examined for randomness and standard strategies for dealing with missing data will be employed as specified in the Statistical Analysis section.

This definition of the Estimand adheres to the modified intention to treat (mITT) principle and will be complemented by the mITT trial analysis.

#### 4.8.2 Secondary outcomes

##### Efficacy

- mRS outcome at 3 months across the full ordinal scale;
- Proportion of participants achieving unassisted walking 50m within 3 months;
- Time (days) to achieving unassisted walking 50m within 3 months;
- Walking speed (10mwt) m/sec at 3 months;
- Quality of life at 3 months (EQ-5D)

##### Safety

- Proportion of participants with mortality at 3 months
- Proportion of participants with mortality at 14 days
- Number and severity of complications (Serious Adverse Events) at 14 days and 3 months post randomisation. We will classify complications as ‘immobility related’ and ‘stroke related’ and examine each class of complications separately.

##### Cost-effectiveness and cost-utility

- Analysis plan for cost-effectiveness and cost-utility analysis will be presented in a separate document.

#### 4.8.3 Tertiary and exploratory outcomes

- Length of hospitalisation (acute plus rehabilitation)
- Blood pressure during the first 3 days of mobility training
- Patient centered outcome measures at 3 and 6 months post-randomisation
- Fatigue (FAS) at 3 and 6 months post-randomisation
- Motor Recovery (FMA) baseline to 3 months post-randomisation
- Cognitive function (MoCA) at 3 months post-randomisation
- Mood (HADS, SADQ) at 3 and 6 months post-randomisation
- Activity limitation (Nottingham Extended ADL) at 3 and 6 months post-randomisation

## 6. Statistical analysis

### 6.1 Analysis principles and general considerations

- All outcomes and analyses are prospectively categorized as primary, secondary, or tertiary/exploratory
- Differences in all endpoints between the three experimental arms of the study and the reference arm will be tested in mild and moderate severity strata independently
- Multi-arm design of AVERT-DOSE requires multiple comparisons involving the primary endpoint. As per the relevant FDA Guideline^(9)^ we will not use multiplicity adjustments to strongly control the probability of making at least one type I error across the multiple comparisons of different regiments to the reference arm. Every individual comparison of a given regimen to the reference arm will be conducted at the two-tailed 0.05 level of significance. Unless specifically stated otherwise, all estimates of treatment effects will be presented with 95% confidence intervals (95%CIs)
- No formal adjustments will be undertaken to constrain the overall type I error associated with the secondary, tertiary, and exploratory analyses. Their purpose is to supplement evidence from the primary analysis to more fully characterize the treatment effect. Results from the secondary and exploratory analyses will be interpreted in this context. No respective p-values will be reported in the manuscript.
- Analysis for primary outcome will be conducted within the Estimand framework using the Estimand described earlier in this SAP while preserving the principle that all participants who did not withdraw consent prior to intervention will be analysed within the arm to which they were randomised, utilising pre-specified strategies for handling intercurrent events.
- The analysis strategy for AVERT-DOSE trial is defined as follows:^(10)^
  ○ It is based on a design that aims to collect all outcome data on all randomised subjects
  ○ It includes a main analysis that keeps subjects in their randomised arms, analyzes all available outcome data, and is valid under a named plausible assumption about the missing data (data missing at random)
  ○ It includes sensitivity analyses that consider a range of plausible alternative assumptions that contradict the main assumption about the missing data
  ○ All individuals are included in sensitivity analyses
- There will be no per protocol analysis in the primary paper for this trial.
- Subgroup analyses could be carried out irrespective of whether there is a significant treatment effect on the outcome. Their purpose is to supplement evidence from the primary analysis to help to fully characterize the treatment effect. Results from subgroup analyses will be interpreted in this context. The following subgroups are prespecified as being of interest for the primary analysis:
  ○ Age: <45 year vs 45-64 years vs 65-80 years vs >80 years, Gender: Male vs Female vs Other
  ○ Stroke type (TOAST): Large Artery Atherosclerosis vs Cardio-Embolism vs Small Artery Occlusion vs SOUE vs other
  ○ Reperfusion treatment: None vs Thrombolysis alone vs EVT+/−Thrombolysis
  ○ Time to randomisation (hours): <24 hours vs 24-48 hours
  ○ Recruitment region: Australia vs Asia vs UK/Europe vs South America
  ○ Pre-stroke physical activity: physically inactive vs higher activity^(11)^
  ○ Pre-stroke clinical frailty: Not frail vs vulnerable^(12)^
- Analyses will be conducted using Stata and R statistical software.

### 6.2 Interim analyses

The Independent DSMB reviewed unblinded data. Due to the blinded change in the trial design in early 2025 from two-stage to the single stage trial, no formal interim analyses for efficacy or futility were conducted.

### 6.3 Trial profile

Flow of patients through the study will be displayed in a standard CONSORT diagram. The report will include the number of patients included, withdrawn, lost to follow up, and the number who were analysed.

### 6.4 Participants’ characteristics and baseline comparisons

In order to assess balance, description of the specified baseline characteristics will be presented by stroke severity strata for all arms. Discrete variables will be summarized as frequencies and percentages. Unless otherwise indicated in the tables, percentages will be calculated according to the number of patients for whom data are available. If there are more than 5% missing values, the denominator will be added in a footnote in the corresponding summary table. Continuous variables will be summarized by use of either mean and standard deviation (SD) or median and interquartile range (IQR). Durations and time intervals will be summarized by medians and IQRs.

### 6.5 Primary outcome: favourable outcome (Independence [little or no disability], mRS0-2) at 3 months

#### Outcome measure

The primary measure of outcome will occur at 3 months post randomisation. The outcome is measured using the mRS.

#### Statistical Hypothesis

For every individual experimental regimen and the reference arm, the set of statistical hypotheses is p(experimental)=p(reference) versus p(experimental) ≠ p(reference), where p(experimental) is the proportion of participants with favourable outcome in the experimental arm and p(reference) is the proportion of participants with favourable outcome in the reference arm.

#### Treatment of missing values

Based on blinded monitoring of the amount of missing primary outcome data in the AVERT-DOSE trial, it is anticipated that there will be limited missing primary outcome data. Nevertheless, a small amount of missing data may be inevitable.

Following clinical input from, and discussions with, the AVERT-DOSE Management Committee, the individual components of the hierarchical primary outcome are assumed to be missing-at-random (MAR), i.e. it is assumed that the probability of the outcome data being missing may depend on the values of the observed data, but not on the values of the missing data. Important explanatory and auxiliary variables such as age, baseline NIHSS, reperfusion treatment status and stroke classification were collected and will be examined to assess the plausibility of the MAR assumption.

For the primary analysis, missing primary outcome data and covariates required for stratification will be imputed using multiple imputations with chained equations (10 imputations, 30 chains, 10000 rerandomisation iterations) with the subsequent use of Rubin’s rules.

Sensitivity analyses using the Tipping Point method^(13,14)^ will be conducted.

#### Analysis method

The effect estimate for the primary efficacy outcome will be obtained using the binary logistic regression model with treatment arms (dummy coded with the reference arm serving as the reference category) as an independent variable and the 3 months mRS outcome (dichotomized into mRS 0-2 as favourable outcome and mRS 3-6 as non-favourable outcome) as the dependent variable, including baseline stroke severity (NIHSS score) and all the factors used for covariate-adaptive randomisation (age, region (Aus; Asia; UK/Europe; and South America) and hyperacute treatment (neither thrombolysis nor endovascular clot retrieval; Thrombolysis but not endovascular clot retrieval; endovascular clot retrieval with or without thrombolysis) as treatment covariates for adjustment purposes. The treatment effect will be presented as adjusted odds ratio (aOR) and standardized risk difference (sRD) with respective 95% CIs as per FDA Guidance for Industry “Adjusting for Covariates in Randomised Clinical Trials for Drugs and Biological Products”.

Hypothesis testing for the primary analysis will be performed via rerandomisation tests as per FDA guidelines^(15)^. Participants will be repeatedly rerandomised into four arms with all outcomes remaining fixed to ensure that the null hypothesis is true. The adjusted OR will be estimated under each rerandomisation and compared to the observed treatment effect. The p-value will be estimated as the proportion of times that a magnitude of the rerandomised aOR is larger than the observed treatment effect.

10,000 replications will be used for the rerandomisation test. If the aOR and sRD are not estimable for any of these replications (due to e.g. all participants in one stratum being allocated to a single arm upon rerandomisation), further replications will be performed in blocks of 5,000 until at least 10,000 successful replications are completed. Excess replications will be discarded from estimation of treatment effects. The proportion of total replications attempted that were not successful will be reported.

Finally, given the early stopping of the trial, the recruited sample size is likely to result in effect estimates that are less precise than originally anticipated. To maximize the opportunity to draw meaningful conclusions from the trial, for every arm we will estimate the frequentist confidence^(16)^ of the given experimental regimen being superior to the reference arm (Conf(sRD>0)) and the frequentist confidence that the given experimental regimen not providing clinically meaningful benefit compared to the reference arm (Conf(sRD<0.05)). The threshold for clinically meaningful benefit of 0.05 (NNT=20) has been chosen by the AVERT-DOSE management committee based on the outcomes of the survey of a multidisciplinary group of clinicians (neurologists and physiotherapists) and people with lived experience of stroke. The survey participants were asked the following question: “In your opinion, how many more patients out of 100 would need to achieve a good outcome (mRS 0–2) with a mobility intervention compared to the control group, for the mobility intervention to be considered meaningful?”

### 6.6 Secondary efficacy outcome: ordinal shift across full mRS at 3 months

#### Outcome measure

This measure of outcome will occur at 3 months post randomisation. The outcome is measured using the mRS.

#### Statistical Hypothesis

For every individual experimental regimen and the reference arm, the set of statistical hypotheses is adjusted common OR=1 versus adjusted common OR ≠ 1.

#### Treatment of missing values

As per the primary outcome.

#### Analysis method

The effect estimate for this secondary efficacy outcome will be obtained using the ordinal logistic regression model with treatment arms (dummy coded with the reference arm serving as the reference category) as an independent variable and the 3 months mRS outcome across full ordinal scale as the dependent variable, including baseline stroke severity (NIHSS score) and all the factors used for covariate-adaptive randomisation (age, region (Aus; Asia; UK/Europe; and South America) and hyperacute treatment (neither thrombolysis nor endovascular clot retrieval; Thrombolysis but not endovascular clot retrieval; endovascular clot retrieval with or without thrombolysis) as treatment covariates for adjustment purposes. The treatment effect will be presented as adjusted common odds ratio (acOR) with respective 95% CIs subject to the condition that all the aOR point estimates across individual binary cut-points for mRS categories are either greater than, or less than 1. Otherwise, stratified generalized Odds Ratios (WinOdds) will be estimated and presented with respective 95%CI.^(17)^

Hypothesis testing for this analysis will be conducted as per primary analysis.

### 6.7 Secondary efficacy outcome: achieving unassisted walking 50m within 3 months

#### Outcome measure

This measure of outcome will occur at 3 months post randomisation.

#### Statistical Hypothesis

For every individual experimental regimen and the reference arm, the set of statistical hypotheses is p(experimental)=p(reference) versus p(experimental) ≠ p(reference), where p(experimental) is the proportion of participants who achieved unassisted walking within 3 months in the experimental arm and p(reference) is the proportion of participants who achieved unassisted walking within 3 months in the reference arm.

#### Treatment of missing values

Complete case analysis will be undertaken.

#### Analysis method

The effect estimate for this outcome will be obtained using the binary logistic regression model with treatment arms (dummy coded with the reference arm serving as the reference category) as an independent variable and achieving unassisted walking within 3 months (dichotomized as yes vs no) as the dependent variable, including baseline stroke severity (NIHSS score) and all the factors used for covariate-adaptive randomisation (age, region (Aus; Asia; UK/Europe; and South America) and hyperacute treatment (neither thrombolysis nor endovascular clot retrieval; Thrombolysis but not endovascular clot retrieval; endovascular clot retrieval with or without thrombolysis) as treatment covariates for adjustment purposes. The treatment effect will be presented as adjusted odds ratio (aOR) and standardized risk difference (sRD) with respective 95% CIs as per FDA Guidance for Industry “Adjusting for Covariates in Randomised Clinical Trials for Drugs and Biological Products”.

### 6.8 Secondary efficacy outcome: time to unassisted walking 50m within 3 months

#### Outcome measure

Time (days) until achieving the outcome of unassisted walking within 3 months post randomisation.

#### Statistical Hypothesis

For every individual experimental regimen and the reference arm, the set of statistical hypotheses is cause-specific adjusted HR=1 versus cause-specific adjusted HR ≠ 1.

#### Treatment of missing values

Complete case analysis will be undertaken.

#### Analysis method

The number of days from stroke onset to walking 50 m unassisted will be used as the time variable for participants who walked, including those who walked and subsequently died within the 3-month period. The number of days from stroke onset to death will be used as the time variable for participants who died and did not walk within the 3-month period. The censoring time of 3 months (range of 89–92 days depending on month of recruitment) will be allocated to participants who did not walk and did not die within 3 months.

Since the event of death provides informative censoring for walking, that is, participants who died are not expected to achieve walking after the event of death, death is considered as a competing risk to the event of walking. Under the circumstances of informative censoring, the use of standard Cox proportional hazards regression is not appropriate and cause-specific modelling will be undertaken.

Subject to the validity of proportional hazards assumption, the effect estimate for this outcome will be obtained using cause-specific competing risk Cox proportional hazards model for days to unassisted walking, with death as the competing risk, treatment arms (dummy coded with the reference arm serving as the reference category) as an independent variable and baseline stroke severity (NIHSS score) and all the factors used for covariate-adaptive randomisation (age, region (Aus; Asia; UK/Europe; and South America) and hyperacute treatment (neither thrombolysis nor endovascular clot retrieval; Thrombolysis but not endovascular clot retrieval; endovascular clot retrieval with or without thrombolysis) as treatment covariates for adjustment purposes. The treatment effect will be presented as adjusted cause-specific Hazard Ratio (acHR) with respective 95% CIs.

### 6.9 Secondary efficacy outcome: walking speed for 10 meters walking test at 3 months

#### Outcome measure

This measure of outcome (m/sec) will occur at 3 months post randomisation.

#### Statistical Hypothesis

For every individual experimental regimen and the reference arm, the set of statistical hypotheses is that the median speed in experimental arm is equal to the median speed in reference arm versus that the median speed in experimental arm is not equal to the median speed in reference arm.

#### Treatment of missing values

Complete case analysis will be undertaken.

#### Analysis method

The effect estimate for this outcome will be obtained using median regression model with treatment arms (dummy coded with the reference arm serving as the reference category) as an independent variable and the walking speed value as the dependent variable, including baseline stroke severity (NIHSS score) and all the factors used for covariate-adaptive randomisation (age, region (Aus; Asia; UK/Europe; and South America) and hyperacute treatment (neither thrombolysis nor endovascular clot retrieval; Thrombolysis but not endovascular clot retrieval; endovascular clot retrieval with or without thrombolysis) as treatment covariates for adjustment purposes. The treatment effect will be presented as adjusted difference in median walking speed value with the corresponding 95% CIs. We will also present respective effect estimates for the 25^th^ and 75^th^ percentile of the walking speed distributions.

### 6.10 Secondary efficacy outcome: Health related quality of life

#### Outcome measure

This secondary measure of outcome is measured at 3 months post-randomisation using the EuroQOL EQ-5D-5L^(18)^, a widely accepted, validated measure. It is a five-dimension scale where each item is scored from 1 to 5. For each dimension (mobility, self-care, usual activities, pain/discomfort, anxiety/depression), respondents are asked to describe their levels of health problems as one of five levels: none (1), slight (2), moderate (3), severe (4) or extreme/enable to perform (5). Thus, the descriptive system is able to identify 3125 unique health states.

A unique health state is defined by combining 1 level from each of the 5 dimensions. Each state is referred to in terms of a 5-digit code. For example, state 11111 indicates no problems on any of the 5 dimensions, while state 12345 indicates no problems with mobility, slight problems with washing or dressing, moderate problems with doing usual activities, severe pain or discomfort and extreme anxiety or depression.

The EQ5D5L measure also includes a 20cm vertical visual analogue scale (VAS). The patient is asked to mark an X on the scale and write the corresponding number in the box to indicate how their health is today. The extremes of the scale are defined as ‘the best health you can imagine’ and ‘the worst health you can imagine’, scoring 100 to 0 respectively. For the VAS, higher values are indicative of better health.

#### Statistical Hypothesis

For every individual experimental regimen and the reference arm, the set of statistical hypotheses is that the median EQ5D5L VAS score in experimental arm is not different to the median EQ5D5L VAS scores in reference arm (null) vs that there is a difference between arms in the median EQ5D5L VAS scores.

#### Treatment of missing values

Complete case analysis will be undertaken.

#### Analysis method

The effect estimate for this outcome will be obtained using median regression model with treatment arms (dummy coded with the reference arm serving as the reference category) as an independent variable and EQ5D5L VAS scores as the dependent variable, including baseline stroke severity (NIHSS score) and all the factors used for covariate-adaptive randomisation (age, region (Aus; Asia; UK/Europe; and South America) and hyperacute treatment (neither thrombolysis nor endovascular clot retrieval; Thrombolysis but not endovascular clot retrieval; endovascular clot retrieval with or without thrombolysis) as treatment covariates for adjustment purposes. The treatment effect will be presented as adjusted difference in median EQ5D5L VAS value with the corresponding 95% CIs. We will also present respective effect estimates for the 25^th^ and 75^th^ percentile of the EQ5D5L VAS distributions.

The analysis of EQ5D5L utility scores will be undertaken as a part of economic analysis and will be detailed in a separate document.

### 6.11 Secondary safety outcome: mortality within 3 months

#### Outcome measure

This measure of outcome will occur at 3 months post randomisation.

#### Statistical Hypothesis

For every individual experimental regimen and the reference arm, the set of statistical hypotheses is p(experimental)=p(reference) versus p(experimental) ≠ p(reference), where p(experimental) is the proportion of participants with mortality events within 3 months in the experimental arm and p(reference) is the proportion of participants with mortality events within 3 months in the reference arm.

#### Treatment of missing values

As per the primary outcome

#### Analysis method

The effect estimate for this outcome will be obtained using the binary logistic regression model with treatment arms (dummy coded with the reference arm serving as the reference category) as an independent variable and mortality within 3 months (dichotomized as yes vs no) as the dependent variable, including baseline stroke severity (NIHSS score) and all the factors used for covariate-adaptive randomisation (age, region (Aus; Asia; UK/Europe; and South America) and hyperacute treatment (neither thrombolysis nor endovascular clot retrieval; Thrombolysis but not endovascular clot retrieval; endovascular clot retrieval with or without thrombolysis) as treatment covariates for adjustment purposes. The treatment effect will be presented as adjusted odds ratio (aOR) and standardized risk difference (sRD) with respective 95% CIs as per FDA Guidance for Industry “Adjusting for Covariates in Randomised Clinical Trials for Drugs and Biological Products”.

### 6.12 Secondary safety outcome: mortality within 14 days

#### Outcome measure

This measure of outcome will occur at 14 days post randomisation.

#### Statistical Hypothesis

For every individual experimental regimen and the reference arm, the set of statistical hypotheses is p(experimental)=p(reference) versus p(experimental) ≠ p(reference), where p(experimental) is the proportion of participants with mortality events within 14 days in the experimental arm and p(reference) is the proportion of participants with mortality within 14 days in the reference arm.

#### Treatment of missing values

As per the primary outcome

#### Analysis method

The effect estimate for this outcome will be obtained using the binary logistic regression model with treatment arms (dummy coded with the reference arm serving as the reference category) as an independent variable and mortality within 3 months (dichotomized as yes vs no) as the dependent variable, including baseline stroke severity (NIHSS score) and all the factors used for covariate-adaptive randomisation (age, region (Aus; Asia; UK/Europe; and South America) and hyperacute treatment (neither thrombolysis nor endovascular clot retrieval; Thrombolysis but not endovascular clot retrieval; endovascular clot retrieval with or without thrombolysis) as treatment covariates for adjustment purposes. The treatment effect will be presented as adjusted odds ratio (aOR) and standardized risk difference (sRD) with respective 95% CIs as per FDA Guidance for Industry “Adjusting for Covariates in Randomised Clinical Trials for Drugs and Biological Products”.

### 6.13 Secondary safety outcome: number of serious adverse events (SAEs) within 3 months

#### Outcome measure

This measure of outcome will occur at 3 months post randomisation.

#### Statistical Hypothesis

For every individual experimental regimen and the reference arm, the set of statistical hypotheses is that the expected per participant count of serious adverse events within 3 months in experimental arm is not different to that in the reference arm vs that the expected per participant count of serious adverse events within 3 months in experimental arm is different to that in the reference arm.

#### Treatment of missing values

Complete case analysis will be undertaken.

#### Analysis method

The effect estimate for this outcome will be obtained using Poisson or negative binomial regression model (depending on the validity of distributional assumptions) with treatment arms (dummy coded with the reference arm serving as the reference category) as an independent variable and the SAE count within 3 months (dichotomized as yes vs no) as the dependent variable, including baseline stroke severity (NIHSS score) and all the factors used for covariate-adaptive randomisation (age, region (Aus; Asia; UK/Europe; and South America) and hyperacute treatment (neither thrombolysis nor endovascular clot retrieval; Thrombolysis but not endovascular clot retrieval; endovascular clot retrieval with or without thrombolysis) as treatment covariates for adjustment purposes. The treatment effect will be presented as adjusted Incidence Rate ratio (aIRR).

### 6.14 Secondary safety outcome: number of serious adverse events (SAEs) within 14 days

#### Outcome measure

This measure of outcome will occur at 14 days post randomisation.

#### Statistical Hypothesis

For every individual experimental regimen and the reference arm, the set of statistical hypotheses is that the expected per participant count of serious adverse events within 14 days in experimental arm is not different to that in the reference arm vs that the expected per participant count of serious adverse events within 14 days in experimental arm is different to that in the reference arm.

#### Treatment of missing values

Complete case analysis will be undertaken.

#### Analysis method

The effect estimate for this outcome will be obtained using Poisson or negative binomial regression model (depending on the validity of distributional assumptions) with treatment arms (dummy coded with the reference arm serving as the reference category) as an independent variable and the SAE count within 14 days (dichotomized as yes vs no) as the dependent variable, including baseline stroke severity (NIHSS score) and all the factors used for covariate-adaptive randomisation (age, region (Aus; Asia; UK/Europe; and South America) and hyperacute treatment (neither thrombolysis nor endovascular clot retrieval; Thrombolysis but not endovascular clot retrieval; endovascular clot retrieval with or without thrombolysis) as treatment covariates for adjustment purposes. The treatment effect will be presented as adjusted Incidence Rate ratio (aIRR).

### 6.15 Tertiary and Exploratory outcomes (some or all of these outcomes may be reported in publications subsequent to primary paper)

Analyses for all the tertiary and exploratory outcomes will be conducted with the aim to estimate the magnitude of respective effect sizes and related uncertainty in the form of the 95%CI. Effect sizes will be presented as stratified/adjusted risk differences and/or odds ratios for the pre-specified dichotomous and categorical outcomes and between-arm differences in the median, 25^th^ percentile and 75^th^ percentile of respective distributions for the pre-specified time metrics.

### 6.16 COVID-19 pandemic related analyses

Due to very small numbers of participants recruited *prior* to the COVID-19 pandemic, the impact of COVID-19 on treatment effect cannot be assessed.

### 6.17 Tables and figures for the main paper

The proposed tables and figures for the main results are presented at the end of this SAP document, together with several future paper tables.

Manuscript Table 1 will report key baseline characteristics of participants by stroke severity strata and by treatment group. Table 2 will report key summary data about the timing, frequency, intensity and amount of training provided by group and stratum. Table 3 will report the primary and key secondary outcomes, and include length of hospital stay, and Table 4 will report safety outcomes. Figure 1 will be the CONSORT diagram. Figure 2 will report “Grotta bars” stacked bar chart of each grade on the mRS in each treatment arm by treatment arm separately for two stroke severity strata. Figure 3 will be a forest plot for the pre-specified subgroup analyses for the primary outcome separately for the two stroke severity strata. Supplemental Figure 1 will include the survival analysis for time to walking.

## Data Availability

All data produced in the present study are available upon reasonable request to the authors

## 2. Abbreviations

ADL: Activities of Daily Living
AE: Adverse Event
aIRR: adjusted Incidence Rate Ratio (aIRR)
aOR: adjusted Odds Ratio
acOR: adjusted common Odds Ratio
acHR: Adjusted cause specific Hazard Ratio
AVERT: Phase 3 randomised controlled, blinded outcome trial of very early mobilisation versus standard care in acute stroke patients.^1^
CFS: Clinical Frailty Scale
CARA: Covariate-Adjusted, Response-Adaptive
CRIq: Cognitive Reserve Index questionnaire
DSMB: Data Safety and Monitoring Committee
ECR: Endovascular Clot Retrieval
eCRF: electronic Case Report Form
EQ-5D-5L: EuroQol, 5 dimensions, 5 levels questionnaire
FAC: Functional Ambulation Classification
FAS: Fatigue Assessment Scale
FDA: Food and Drug Administration
FMA: Fugl-Meyer Assessment
FMA-UL: Fugl-Meyer Assessment of Upper Limb
FMA-LL: Fugl-Meyer Assessment of Lower Limb
GCP: Good Clinical Practice
HADS: Hospital Anxiety and Depression Scale
HR: Hazard Ratio
ICH: International Conference on Harmonisation
ID: Identification
IME: Important Medical Event
IQR: Inter-Quartile Range
MAMS: Multi-Arm Multi-Stage
mITT: Modified Intention To Treat
mmHg: millimetres of Mercury
MoCA: Montreal Cognitive Assessment
MRI: Magnetic Resonance Imaging
mRS: modified Rankin Scale
MSAS: Mobility Scale for Acute Stroke
NEADL: Nottingham Extended ADL scale
NHMRC: National Health and Medical Research Council
NIHSS: National Institute of Health Stroke Scale
OR: Odds Ratio
PHP: Hypertext Preprocessor
SADQ-H 10: Stroke Aphasic Depression Questionnaire – Hospital Version 10 items
SAE: Serious Adverse Event
SBP: Systolic Blood Pressure
SD: Standard Deviation
SGPALS: Saltin-Grimby Physical Activity Level Scale
sRD: standardized Risk Difference
SRRR: Stroke Recovery and Roundtable
TOAST: Trial of ORG 10172 in Acute Stroke Treatment. (Stroke subtype Classification)
UK: United Kingdom
VAS: Visual Analogue Scale
10MWT: Ten Metre Walk Test

## 8. Manuscript Tables and Figures

**MANUSCRIPT TABLE 1:**
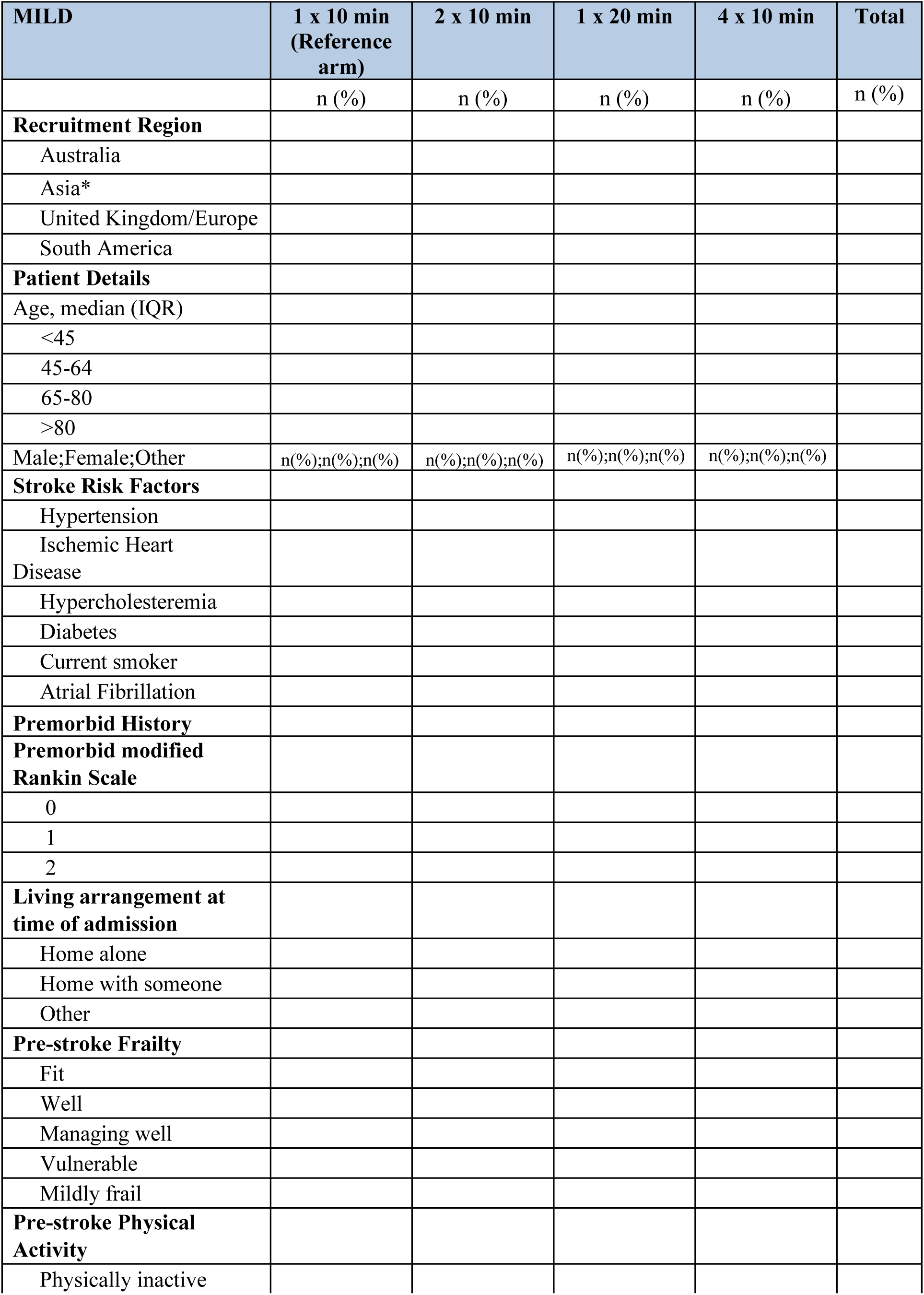
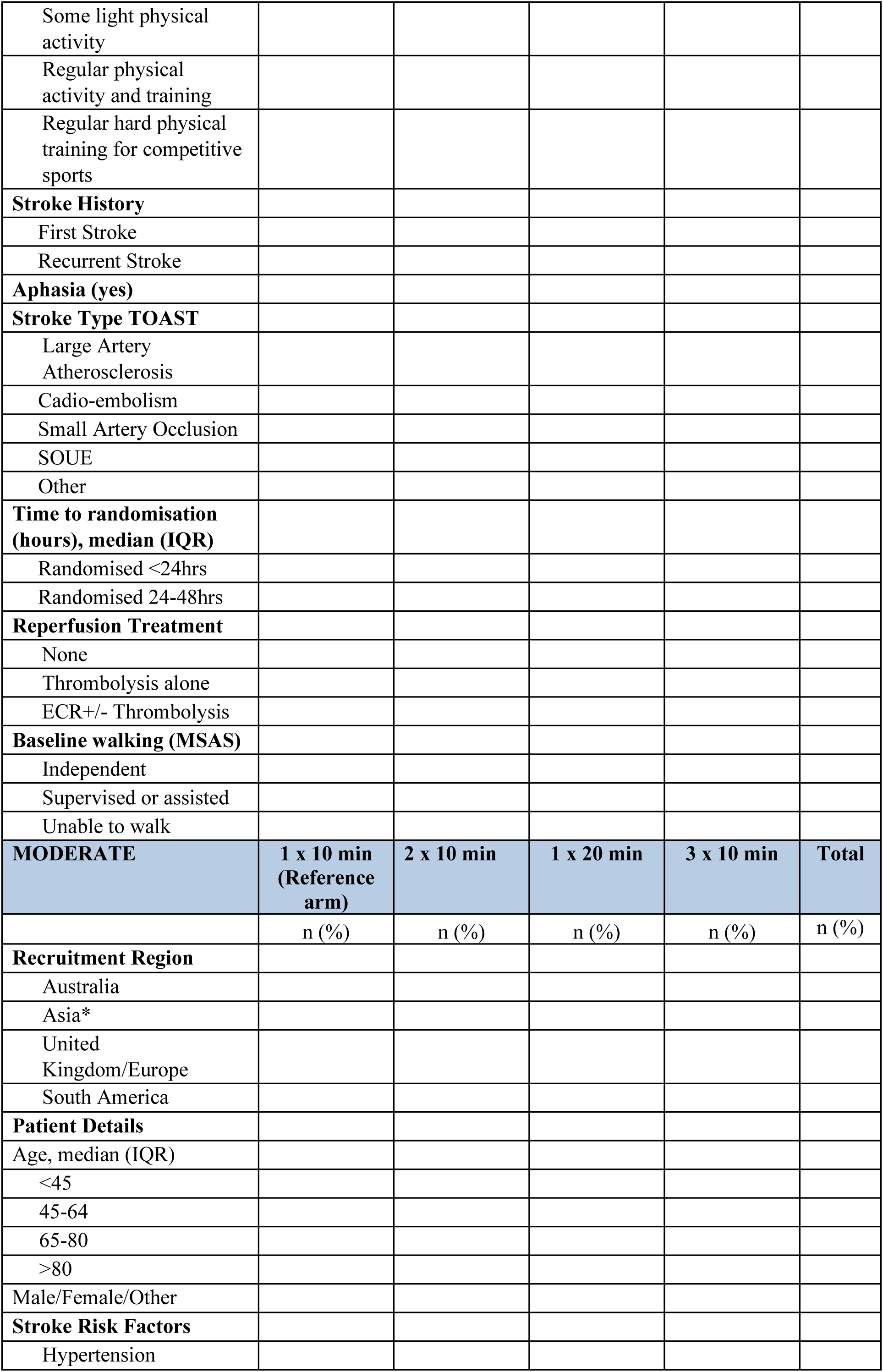
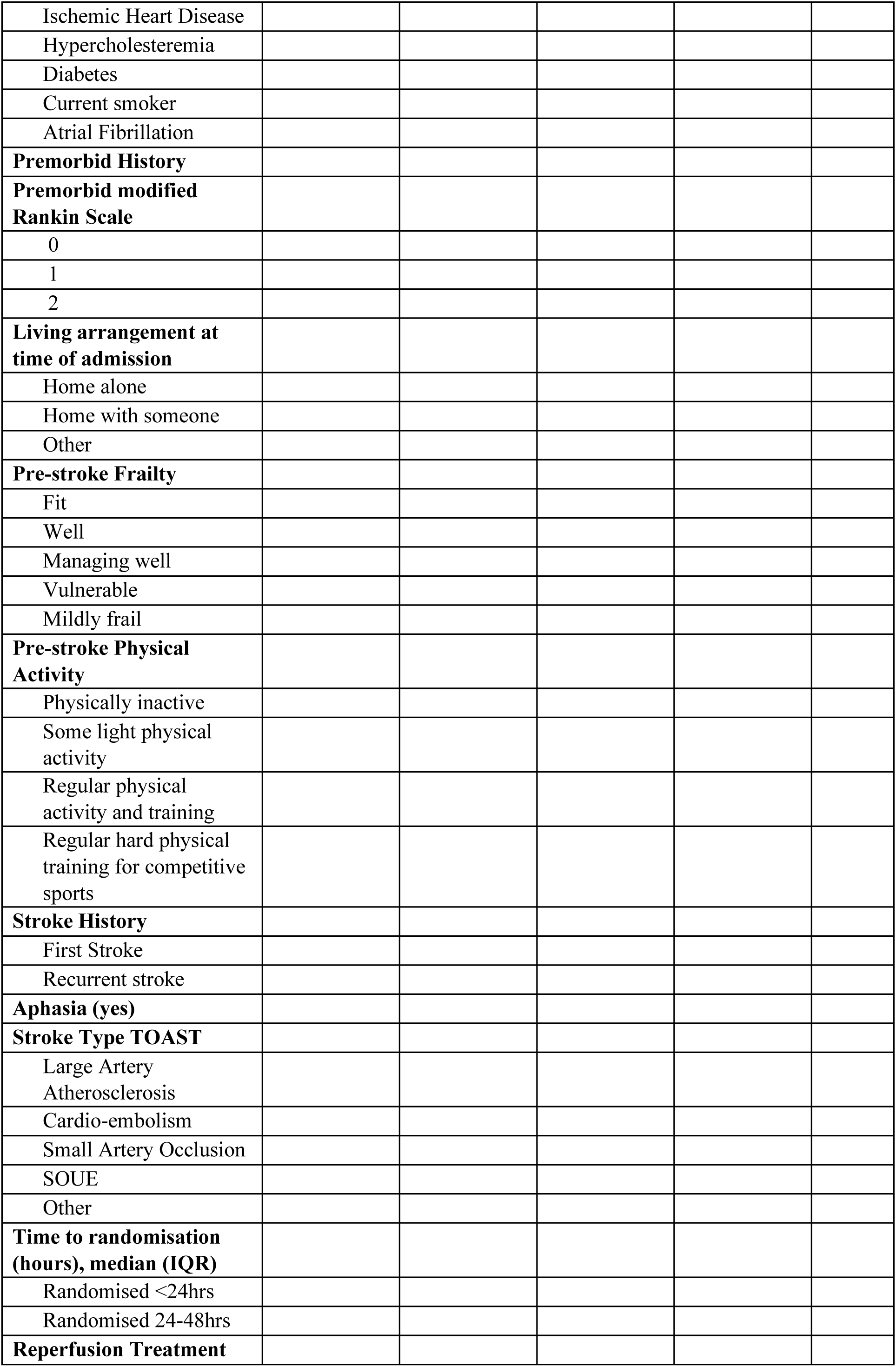
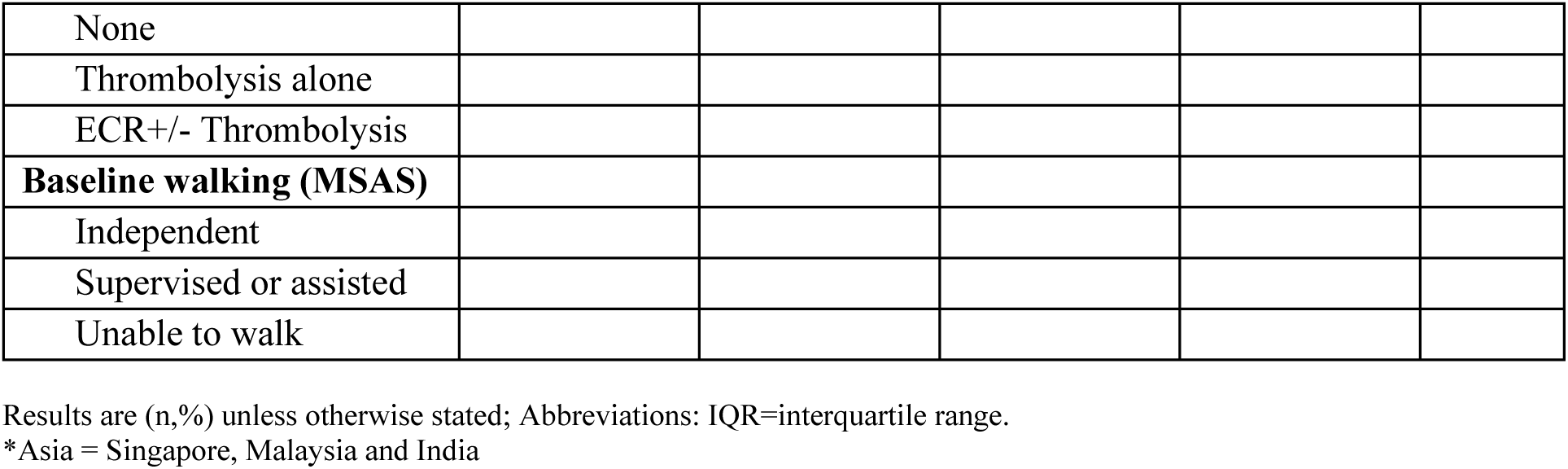
Baseline characteristics of recruited patients: Mild and Moderate Strata (n=)

**MANUSCRIPT TABLE 2:**
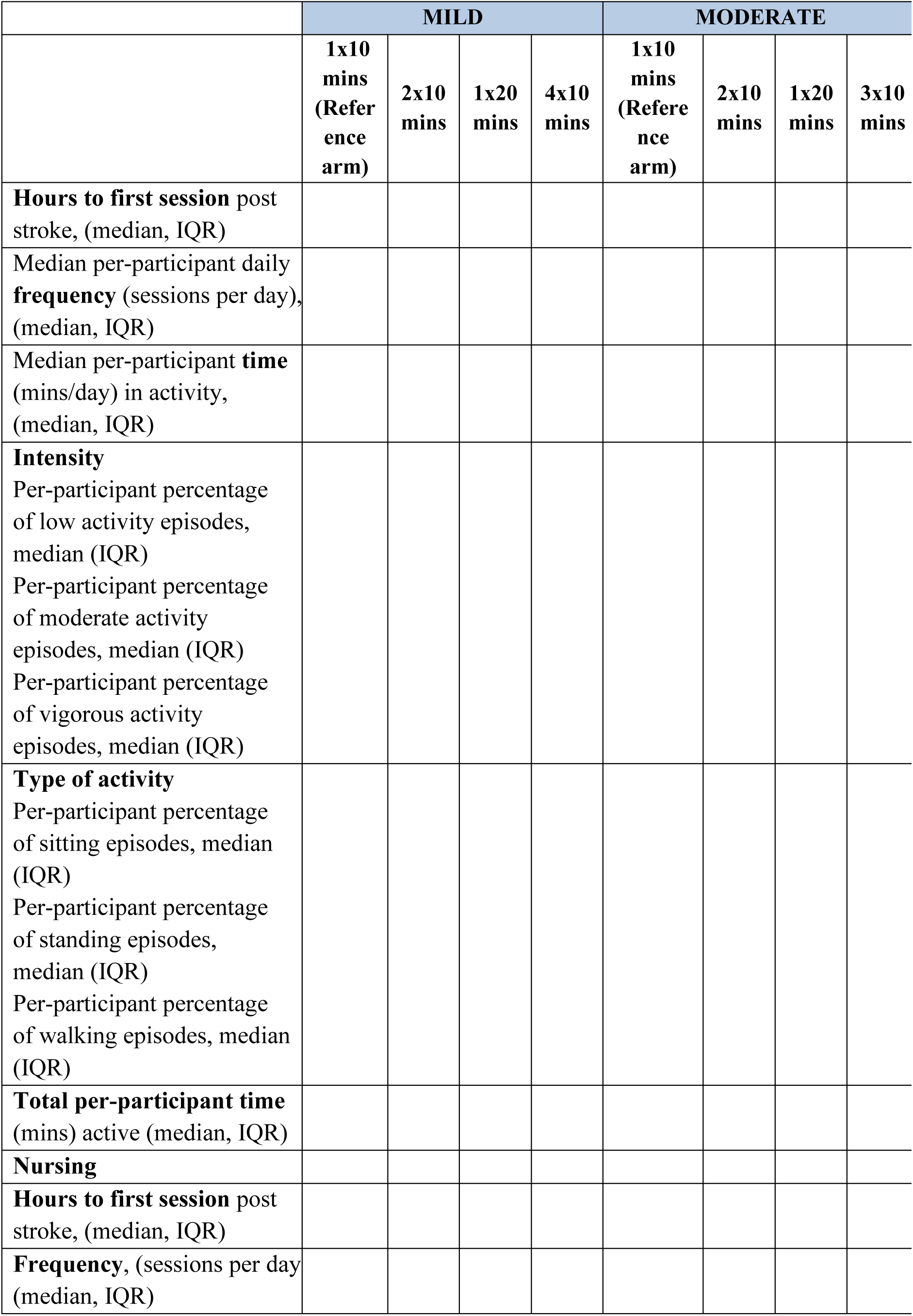
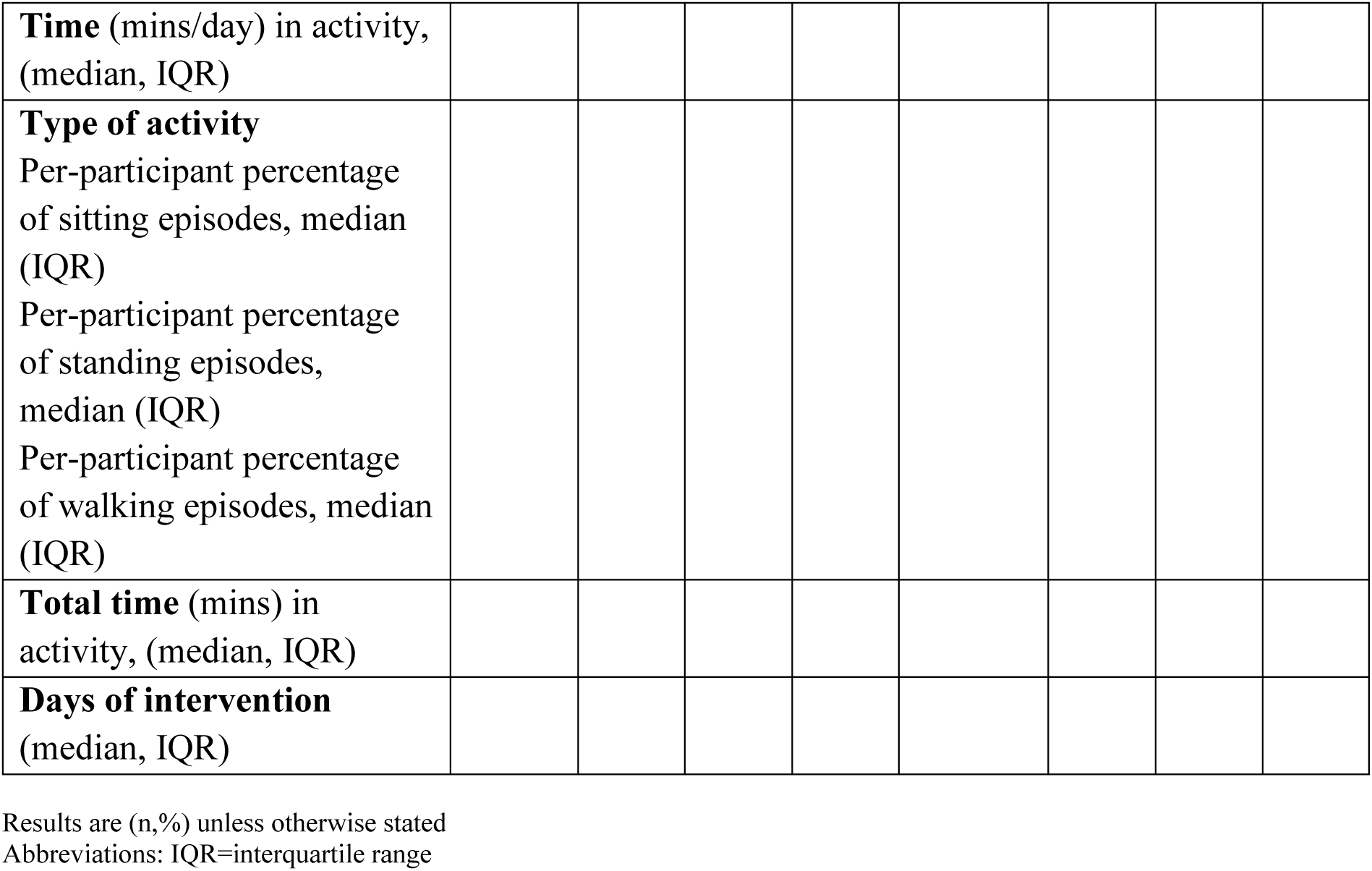
Intervention characteristics: Mild and Moderate Strata.

**MANUSCRIPT TABLE 3:**
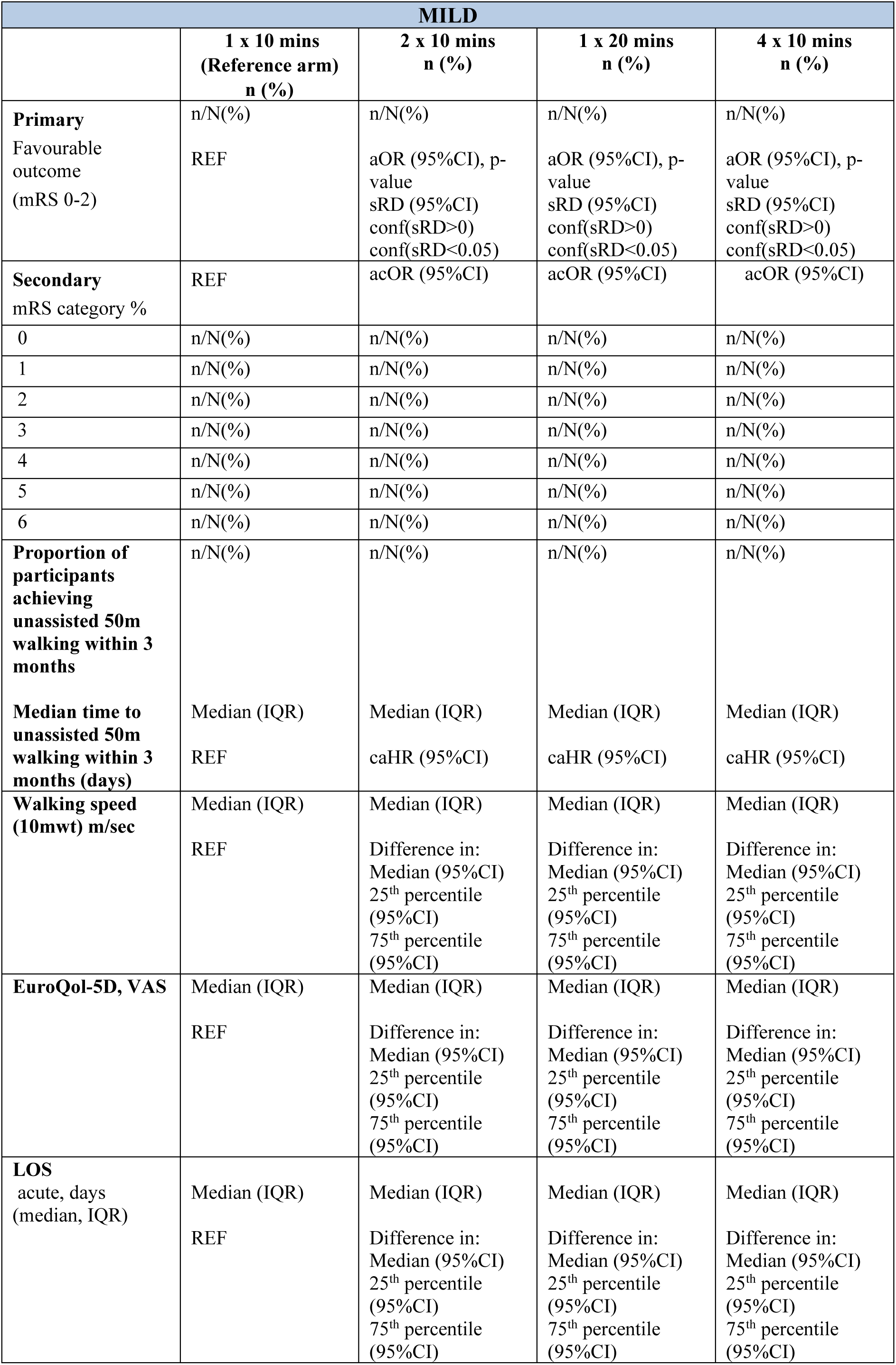
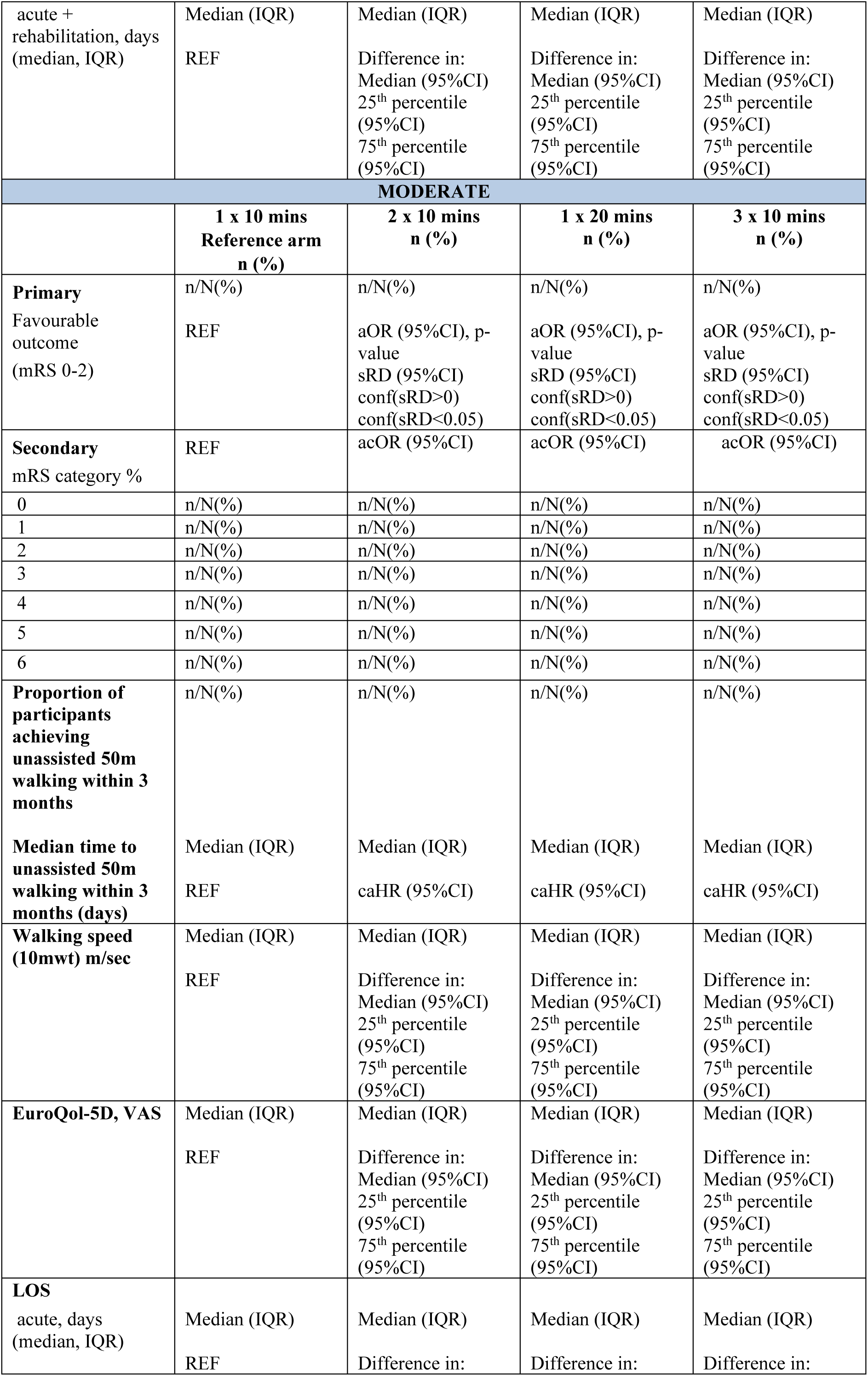
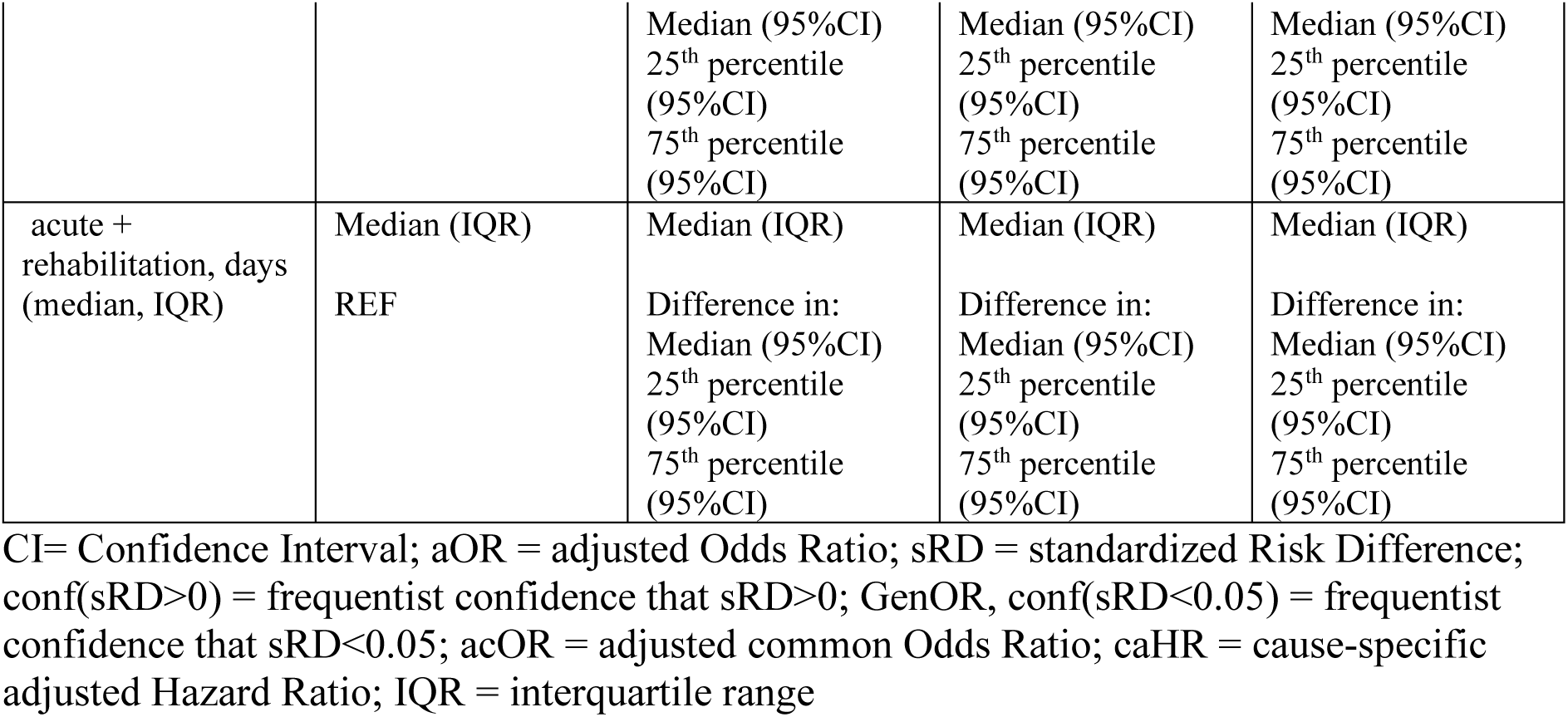
Primary and Secondary Outcomes at 3 months.

| **MILD** |  |  |  |  |
| --- | --- | --- | --- | --- |
|  | **1 x 10 mins**<br>**(Reference arm)**<br>**n (%)** | **2 x 10 mins**<br>**n (%)** | **1 x 20 mins**<br>**n (%)** | **4 x 10 mins**<br>**n (%)** |
| **Primary**<br>Favourable outcome<br>(mRS 0-2) | n/N(%)<br>REF | n/N(%)<br>aOR (95%CI), p-value<br>sRD (95%CI)<br>conf(sRD>0)<br>conf(sRD<0.05) | n/N(%)<br>aOR (95%CI), p-value<br>sRD (95%CI)<br>conf(sRD>0)<br>conf(sRD<0.05) | n/N(%)<br>aOR (95%CI), p-value<br>sRD (95%CI)<br>conf(sRD>0)<br>conf(sRD<0.05) |
| **Secondary**<br>mRS category % | REF | acOR (95%CI) | acOR (95%CI) | acOR (95%CI) |
| 0 | n/N(%) | n/N(%) | n/N(%) | n/N(%) |
| 1 | n/N(%) | n/N(%) | n/N(%) | n/N(%) |
| 2 | n/N(%) | n/N(%) | n/N(%) | n/N(%) |
| 3 | n/N(%) | n/N(%) | n/N(%) | n/N(%) |
| 4 | n/N(%) | n/N(%) | n/N(%) | n/N(%) |
| 5 | n/N(%) | n/N(%) | n/N(%) | n/N(%) |
| 6 | n/N(%) | n/N(%) | n/N(%) | n/N(%) |
| **Proportion of participants achieving unassisted 50m walking within 3 months** | n/N(%) | n/N(%) | n/N(%) | n/N(%) |
| **Median time to unassisted 50m walking within 3 months (days)** | Median (IQR)<br>REF | Median (IQR)<br>caHR (95%CI) | Median (IQR)<br>caHR (95%CI) | Median (IQR)<br>caHR (95%CI) |
| **Walking speed (10mwt) m/sec** | Median (IQR)<br>REF | Median (IQR)<br>Difference in:<br>Median (95%CI)<br>25<sup>th</sup> percentile (95%CI)<br>75<sup>th</sup> percentile (95%CI) | Median (IQR)<br>Difference in:<br>Median (95%CI)<br>25<sup>th</sup> percentile (95%CI)<br>75<sup>th</sup> percentile (95%CI) | Median (IQR)<br>Difference in:<br>Median (95%CI)<br>25<sup>th</sup> percentile (95%CI)<br>75<sup>th</sup> percentile (95%CI) |
| **EuroQol-5D, VAS** | Median (IQR)<br>REF | Median (IQR)<br>Difference in:<br>Median (95%CI)<br>25<sup>th</sup> percentile (95%CI)<br>75<sup>th</sup> percentile (95%CI) | Median (IQR)<br>Difference in:<br>Median (95%CI)<br>25<sup>th</sup> percentile (95%CI)<br>75<sup>th</sup> percentile (95%CI) | Median (IQR)<br>Difference in:<br>Median (95%CI)<br>25<sup>th</sup> percentile (95%CI)<br>75<sup>th</sup> percentile (95%CI) |
| **LOS**<br>acute, days<br>(median, IQR) | Median (IQR)<br>REF | Median (IQR)<br>Difference in:<br>Median (95%CI)<br>25<sup>th</sup> percentile (95%CI)<br>75<sup>th</sup> percentile (95%CI) | Median (IQR)<br>Difference in:<br>Median (95%CI)<br>25<sup>th</sup> percentile (95%CI)<br>75<sup>th</sup> percentile (95%CI) | Median (IQR)<br>Difference in:<br>Median (95%CI)<br>25<sup>th</sup> percentile (95%CI)<br>75<sup>th</sup> percentile (95%CI) |
| acute + rehabilitation, days (median, IQR) | Median (IQR)<br>REF | Median (IQR)<br>Difference in: Median (95%CI)<br>25<sup>th</sup> percentile (95%CI)<br>75<sup>th</sup> percentile (95%CI) | Median (IQR)<br>Difference in: Median (95%CI)<br>25<sup>th</sup> percentile (95%CI)<br>75<sup>th</sup> percentile (95%CI) | Median (IQR)<br>Difference in: Median (95%CI)<br>25<sup>th</sup> percentile (95%CI)<br>75<sup>th</sup> percentile (95%CI) |
| **MODERATE** |  |  |  |  |
|  | **1 x 10 mins**<br>**Reference arm**<br>**n (%)** | **2 x 10 mins**<br>**n (%)** | **1 x 20 mins**<br>**n (%)** | **3 x 10 mins**<br>**n (%)** |
| **Primary**<br>Favourable outcome (mRS 0-2) | n/N(%)<br>REF | n/N(%)<br>aOR (95%CI), p-value<br>sRD (95%CI)<br>conf(sRD>0)<br>conf(sRD<0.05) | n/N(%)<br>aOR (95%CI), p-value<br>sRD (95%CI)<br>conf(sRD>0)<br>conf(sRD<0.05) | n/N(%)<br>aOR (95%CI), p-value<br>sRD (95%CI)<br>conf(sRD>0)<br>conf(sRD<0.05) |
| **Secondary**<br>mRS category % | REF | acOR (95%CI) | acOR (95%CI) | acOR (95%CI) |
| 0 | n/N(%) | n/N(%) | n/N(%) | n/N(%) |
| 1 | n/N(%) | n/N(%) | n/N(%) | n/N(%) |
| 2 | n/N(%) | n/N(%) | n/N(%) | n/N(%) |
| 3 | n/N(%) | n/N(%) | n/N(%) | n/N(%) |
| 4 | n/N(%) | n/N(%) | n/N(%) | n/N(%) |
| 5 | n/N(%) | n/N(%) | n/N(%) | n/N(%) |
| 6 | n/N(%) | n/N(%) | n/N(%) | n/N(%) |
| **Proportion of participants achieving unassisted 50m walking within 3 months** | n/N(%) | n/N(%) | n/N(%) | n/N(%) |
| **Median time to unassisted 50m walking within 3 months (days)** | Median (IQR)<br>REF | Median (IQR)<br>caHR (95%CI) | Median (IQR)<br>caHR (95%CI) | Median (IQR)<br>caHR (95%CI) |
| **Walking speed (10mwt) m/sec** | Median (IQR)<br>REF | Median (IQR)<br>Difference in: Median (95%CI)<br>25<sup>th</sup> percentile (95%CI)<br>75<sup>th</sup> percentile (95%CI) | Median (IQR)<br>Difference in: Median (95%CI)<br>25<sup>th</sup> percentile (95%CI)<br>75<sup>th</sup> percentile (95%CI) | Median (IQR)<br>Difference in: Median (95%CI)<br>25<sup>th</sup> percentile (95%CI)<br>75<sup>th</sup> percentile (95%CI) |
| **EuroQol-5D, VAS** | Median (IQR)<br>REF | Median (IQR)<br>Difference in: Median (95%CI)<br>25<sup>th</sup> percentile (95%CI)<br>75<sup>th</sup> percentile (95%CI) | Median (IQR)<br>Difference in: Median (95%CI)<br>25<sup>th</sup> percentile (95%CI)<br>75<sup>th</sup> percentile (95%CI) | Median (IQR)<br>Difference in: Median (95%CI)<br>25<sup>th</sup> percentile (95%CI)<br>75<sup>th</sup> percentile (95%CI) |
| **LOS**<br>acute, days (median, IQR) | Median (IQR)<br>REF | Median (IQR)<br>Difference in: | Median (IQR)<br>Difference in: | Median (IQR)<br>Difference in: |

|  |  | Median (95%CI)<br>25<sup>th</sup> percentile<br>(95%CI)<br>75<sup>th</sup> percentile<br>(95%CI) | Median (95%CI)<br>25<sup>th</sup> percentile<br>(95%CI)<br>75<sup>th</sup> percentile<br>(95%CI) | Median (95%CI)<br>25<sup>th</sup> percentile<br>(95%CI)<br>75<sup>th</sup> percentile<br>(95%CI) |
| --- | --- | --- | --- | --- |
| acute +<br>rehabilitation, days<br>(median, IQR) | Median (IQR)<br>REF | Median (IQR)<br>Difference in:<br>Median (95%CI)<br>25<sup>th</sup> percentile<br>(95%CI)<br>75<sup>th</sup> percentile<br>(95%CI) | Median (IQR)<br>Difference in:<br>Median (95%CI)<br>25<sup>th</sup> percentile<br>(95%CI)<br>75<sup>th</sup> percentile<br>(95%CI) | Median (IQR)<br>Difference in:<br>Median (95%CI)<br>25<sup>th</sup> percentile<br>(95%CI)<br>75<sup>th</sup> percentile<br>(95%CI) |

CI= Confidence Interval; aOR = adjusted Odds Ratio; sRD = standardized Risk Difference; conf(sRD>0) = frequentist confidence that sRD>0; GenOR, conf(sRD<0.05) = frequentist confidence that sRD<0.05; acOR = adjusted common Odds Ratio; caHR = cause-specific adjusted Hazard Ratio; IQR = interquartile range

**MANUSCRIPT TABLE 4:** Deaths and Serious Adverse Events (SAE) at 14 days and at 3 months post randomisation.

| **MILD** |  |  |  |  |
| --- | --- | --- | --- | --- |
|  | **1 x 10 mins**<br>**(Reference arm)**<br>**n (%)** | **2 x 10**<br>**mins**<br>**n (%)** | **1 x 20**<br>**mins**<br>**n (%)** | **4 x 10**<br>**mins**<br>**n (%)** |
| **Deaths** |  |  |  |  |
| 14 days | n/N(%)<br>REF | n/N(%)<br>aOR<br>(95%CI)<br>sRD<br>(95%CI) | n/N(%)<br>aOR<br>(95%CI)<br>sRD<br>(95%CI) | n/N(%)<br>aOR<br>(95%CI)<br>sRD<br>(95%CI) |
| 3 months | n/N(%)<br>REF | n/N(%)<br>aOR<br>(95%CI)<br>sRD<br>(95%CI) | n/N(%)<br>aOR<br>(95%CI)<br>sRD<br>(95%CI) | n/N(%)<br>aOR<br>(95%CI)<br>sRD<br>(95%CI) |
| **SAEs at 14 days** | REF | aIRR<br>(95%CI) | aIRR<br>(95%CI) | aIRR<br>(95%CI) |
| None | n/N(%) | n/N(%) | n/N(%) | n/N(%) |
| 1 | n/N(%) | n/N(%) | n/N(%) | n/N(%) |
| 2 | n/N(%) | n/N(%) | n/N(%) | n/N(%) |
| 3 | n/N(%) | n/N(%) | n/N(%) | n/N(%) |
| >3 | n/N(%) | n/N(%) | n/N(%) | n/N(%) |
| **SAEs at 3 months** |  |  |  |  |
| None | REF<br>n/N(%) | aIRR<br>(95%CI)<br>n/N(%) | aIRR<br>(95%CI)<br>n/N(%) | aIRR<br>(95%CI)<br>n/N(%) |
| 1 | n/N(%) | n/N(%) | n/N(%) | n/N(%) |
| 2 | n/N(%) | n/N(%) | n/N(%) | n/N(%) |
| 3 | n/N(%) | n/N(%) | n/N(%) | n/N(%) |
| >3 | n/N(%) | n/N(%) | n/N(%) | n/N(%) |
| **Stroke-related complications 14 days** |  |  |  |  |
| None | n/N(%) | n/N(%) | n/N(%) | n/N(%) |
| 1 | n/N(%) | n/N(%) | n/N(%) | n/N(%) |
| 2 | n/N(%) | n/N(%) | n/N(%) | n/N(%) |
| 3 | n/N(%) | n/N(%) | n/N(%) | n/N(%) |
| >3 | n/N(%) | n/N(%) | n/N(%) | n/N(%) |
| **Immobility complications 14 days** |  |  |  |  |
| None | n/N(%) | n/N(%) | n/N(%) | n/N(%) |
| 1 | n/N(%) | n/N(%) | n/N(%) | n/N(%) |
| 2 | n/N(%) | n/N(%) | n/N(%) | n/N(%) |
| 3 | n/N(%) | n/N(%) | n/N(%) | n/N(%) |
| >3 | n/N(%) | n/N(%) | n/N(%) | n/N(%) |
| **MODERATE** |  |  |  |  |
|  | **1 x 10 mins**<br>**(Reference arm)**<br>**n (%)** | **2 x 10**<br>**mins**<br>**n (%)** | **1 x 20**<br>**mins**<br>**n (%)** | **3 x 10**<br>**mins**<br>**n (%)** |
| **Deaths** |  |  |  |  |
| 14 days | n/N(%)<br>REF | n/N(%)<br>aOR<br>(95%CI)<br>sRD<br>(95%CI) | n/N(%)<br>aOR<br>(95%CI)<br>sRD<br>(95%CI) | n/N(%)<br>aOR<br>(95%CI)<br>sRD<br>(95%CI) |
| 3 months | n/N(%)<br>REF | n/N(%)<br>aOR<br>(95%CI)<br>sRD<br>(95%CI) | n/N(%)<br>aOR<br>(95%CI)<br>sRD<br>(95%CI) | n/N(%)<br>aOR<br>(95%CI)<br>sRD<br>(95%CI) |
| **SAEs at 14 days** | REF | aIRR<br>(95%CI) | aIRR<br>(95%CI) | aIRR<br>(95%CI) |
| None | n/N(%) | n/N(%) | n/N(%) | n/N(%) |
| 1 | n/N(%) | n/N(%) | n/N(%) | n/N(%) |
| 2 | n/N(%) | n/N(%) | n/N(%) | n/N(%) |
| 3 | n/N(%) | n/N(%) | n/N(%) | n/N(%) |
| >3 | n/N(%) | n/N(%) | n/N(%) | n/N(%) |
| **SAEs at 3 months** |  |  |  |  |
| None | REF | aIRR<br>(95%CI) | aIRR<br>(95%CI) | aIRR<br>(95%CI) |
| 1 | n/N(%) | n/N(%) | n/N(%) | n/N(%) |
| 2 | n/N(%) | n/N(%) | n/N(%) | n/N(%) |
| 3 | n/N(%) | n/N(%) | n/N(%) | n/N(%) |
| >3 | n/N(%) | n/N(%) | n/N(%) | n/N(%) |
| **Stroke-related complications 3 months** |  |  |  |  |
| None | n/N(%) | n/N(%) | n/N(%) | n/N(%) |
| 1 | n/N(%) | n/N(%) | n/N(%) | n/N(%) |
| 2 | n/N(%) | n/N(%) | n/N(%) | n/N(%) |
| 3 | n/N(%) | n/N(%) | n/N(%) | n/N(%) |
| >3 | n/N(%) | n/N(%) | n/N(%) | n/N(%) |
| **Immobility complications 3 months** |  |  |  |  |
| None | n/N(%) | n/N(%) | n/N(%) | n/N(%) |
| 1 | n/N(%) | n/N(%) | n/N(%) | n/N(%) |
| 2 | n/N(%) | n/N(%) | n/N(%) | n/N(%) |
| 3 | n/N(%) | n/N(%) | n/N(%) | n/N(%) |
| >3 | n/N(%) | n/N(%) | n/N(%) | n/N(%) |

CI= Confidence Interval; aOR = adjusted Odds Ratio; sRD = standardized Risk Difference; aIRR = adjusted Incidence Rate Ratio; IQR = interquartile range

**Manuscript Supplementary Table 1:**
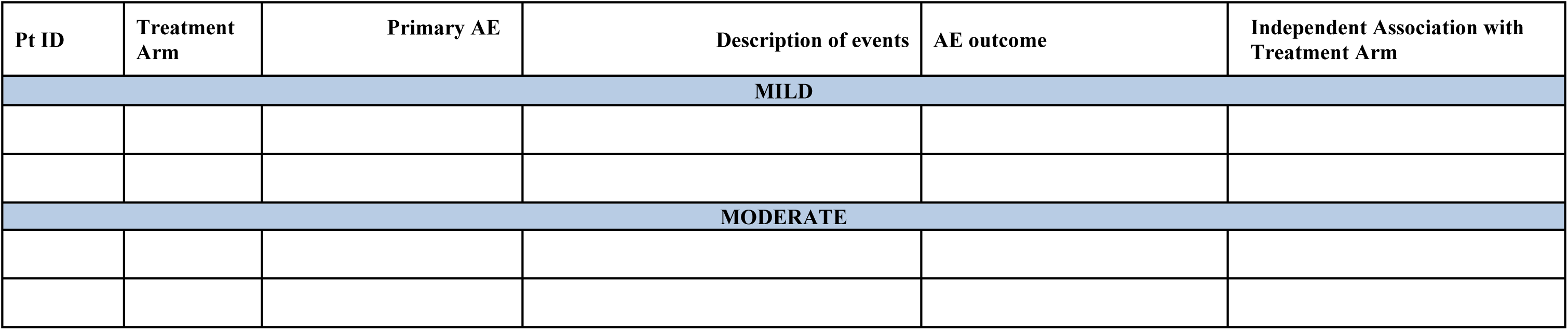
Full List of Serious Adverse Events within 14 days.

**Manuscript Supplementary Table 2:**
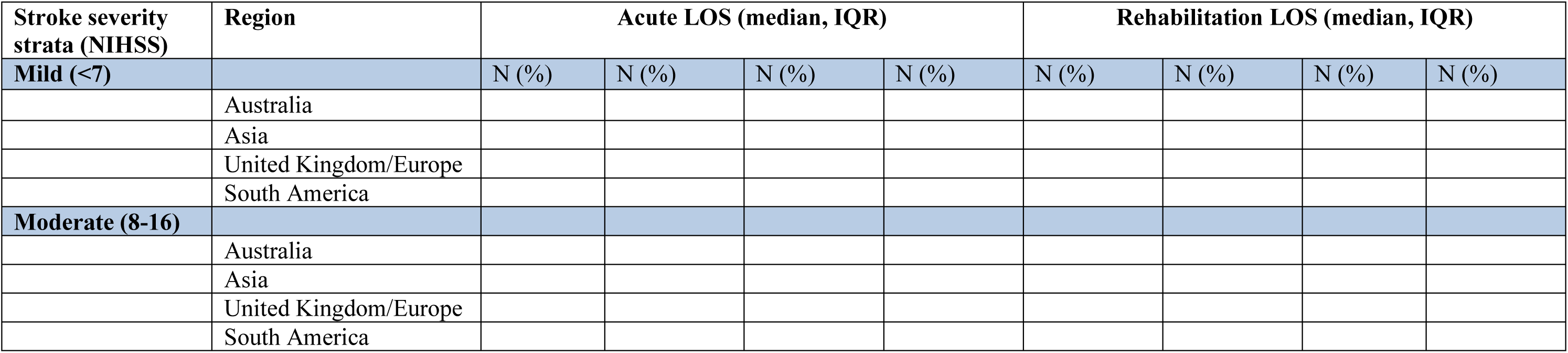
Length of inpatient hospital stay by country.

**Figure 1: Consort Diagram**

**Figure 2: Primary Outcome by stratum (mRS “Grotta” bars by treatment arm)**

**Figure 3: Forest plots for the pre-specified subgroup analyses for the primary outcome**

**Figure S1: The time to unassisted 50m walking plot**.

**Figure S2: Forest plots for the pre-specified subgroup analyses for the selected secondary outcomes (mortality)**.

**Future exploratory papers Table:** Additional baseline characteristics by stratum and group.

| **MILD** | **1 x 10 mins**<br>**(Reference**<br>**arm)** | **2 x 10**<br>**mins** | **1 x 20**<br>**mins** | **4 x 10**<br>**mins** | **Total** |
| --- | --- | --- | --- | --- | --- |
|  | n (%) | n (%) | n (%) | n (%) | n (%) |
| **Cognition** |  |  |  |  |  |
| Cog 4 |  |  |  |  |  |
| Line bisection |  |  |  |  |  |
| Trial Making A |  |  |  |  |  |
| **Fugl-Meyer Assessment (FMA)** |  |  |  |  |  |
| FMA-Upper limb |  |  |  |  |  |
| FMA-Lower limb |  |  |  |  |  |
| **Cognitive reserve Index**<br>**questionnaire** |  |  |  |  |  |
| **10MWT (median IQR) hospital**<br>**discharge** |  |  |  |  |  |
| **MODERATE** | **1 x 10 mins**<br>**(Reference**<br>**arm)** | **2 x 10**<br>**mins** | **1 x 20**<br>**mins** | **3 x 10**<br>**mins** | **Total** |
|  | n (%) | n (%) | n (%) | n (%) | n (%) |
| **Cognition** |  |  |  |  |  |
| Cog 4 |  |  |  |  |  |
| Line bisection |  |  |  |  |  |
| Trial Making A |  |  |  |  |  |
| **Fugl-Meyer Assessment (FMA)** |  |  |  |  |  |
| FMA-Upper limb |  |  |  |  |  |
| FMA-Lower limb |  |  |  |  |  |
| **Cognitive reserve Index**<br>**questionnaire** |  |  |  |  |  |
| **10MWT (median IQR) hospital**<br>**discharge** |  |  |  |  |  |

**Future exploratory paper Table:** Additional 3 month outcomes by stratum and group.

| **MILD** | **1 by 10 mins**<br>**(Reference**<br>**arm)** | **2 by 10**<br>**mins** | **1 by 20**<br>**mins** | **4 by 10**<br>**mins** | **Total** |
| --- | --- | --- | --- | --- | --- |
|  | n (%) | n (%) | n (%) | n (%) | n (%) |
| **Cognitive outcomes** |  |  |  |  |  |
| Cog 4 |  |  |  |  |  |
| Trail Making A* |  |  |  |  |  |
| Line bisection* |  |  |  |  |  |
| MOCA |  |  |  |  |  |
| **Fugl-Meyer Assessment**<br>**(FMA)\*** |  |  |  |  |  |
| FMA-Upper limb |  |  |  |  |  |
| FMA-Lower limb |  |  |  |  |  |
| **Nottingham Extended ADL** |  |  |  |  |  |
| **Change in 10MWT**<br>**Baseline# to 3 months** |  |  |  |  |  |
| **FAC** |  |  |  |  |  |
| Non ambulation |  |  |  |  |  |
| Nonfunctional ambulation |  |  |  |  |  |
| Household ambulation |  |  |  |  |  |
| Surroundings of the house<br>ambulation |  |  |  |  |  |
| Independent community<br>ambulation |  |  |  |  |  |
| Normal ambulation |  |  |  |  |  |
| **Fatigue Assessment Scale** |  |  |  |  |  |
| **Additional Patient Centered**<br>**Outcome Measures for Stroke** |  |  |  |  |  |
| How close are you to a good<br>(=100%) recovery? |  |  |  |  |  |
| Do you still have hope for a good<br>recovery?<br>Yes<br>No |  |  |  |  |  |
| Where you driving prior to<br>your stroke<br>Yes<br>No |  |  |  |  |  |
| Are you currently driving<br>Yes<br>No |  |  |  |  |  |
| **MODERATE** | **1 by 10 mins**<br>**(Reference**<br>**arm)** | **2 by 10**<br>**mins** | **1 by 20**<br>**mins** | **3 by 10**<br>**mins** | **Total** |
|  | n (%) | n (%) | n (%) | n (%) | n (%) |
| **Cognitive outcomes** |  |  |  |  |  |
| Cog 4 |  |  |  |  |  |
| Trail Making A* |  |  |  |  |  |
| Line bisection* |  |  |  |  |  |
| MOCA |  |  |  |  |  |
| **Fugl-Meyer Assessment (FMA)\*** |  |  |  |  |  |
| FMA-Upper limb |  |  |  |  |  |
| FMA-Lower limb |  |  |  |  |  |
| **Nottingham Extended ADL** |  |  |  |  |  |
| **Change in 10MWT**<br>Baseline# to 3 months |  |  |  |  |  |
| **FAC** |  |  |  |  |  |
| Nonambulation |  |  |  |  |  |
| Nonfunctional ambulation |  |  |  |  |  |
| Household ambulation |  |  |  |  |  |
| Surroundings of the house ambulation |  |  |  |  |  |
| Independent community ambulation |  |  |  |  |  |
| Normal ambulation |  |  |  |  |  |
| **Fatigue Assessment Scale** |  |  |  |  |  |
| **Additional Patient Centered Outcome Measures for Stroke** |  |  |  |  |  |
| How close are you to a good (=100%) recovery? |  |  |  |  |  |
| Do you still have hope for a good recovery?<br>Yes<br>No |  |  |  |  |  |
| Where you driving prior to your stroke<br>Yes<br>No |  |  |  |  |  |
| Are you currently driving<br>Yes<br>No |  |  |  |  |  |

#10MWT collected before discharge, FMA Fugl-Meyer Assessment, \*These assessments completed up until protocol amendment 4 (July 2024) removing the need to complete.

## References

1. Pocock SJ and Simon R, Sequential treatment assignment with balancing for prognostic factors in the controlled clinical trial. Biometrics, 1975; 31(1):103–15.

2. Wei LJ, Lachin JM. Properties of the urn randomization in clinical trials. Control Clin Trials. 1988 Dec;9(4):345–64

3. Ning & Huang, Response-adaptive randomization for clinical trials with adjustment for covariate imbalance Statistics in Medicine 2010 Volume 29,17:1761-1860 10.1002/sim.3978

4. The AVERT Trial Collaboration group, Bernhardt J, Langhorne P, Lindley RI, Thrift AG, Ellery F, et al. Efficacy and safety of very early mobilisation within 24 h of stroke onset (AVERT): a randomised controlled trial. Lancet. 2015;386:46–55

5. Mehta CR and Pocock SJ. Adaptive increase in sample size when interim results are promising: a practical guide with examples. Statistics in Medicine, 2011;30(28):3267–84. doi: 10.1002/sim.4102

6. International Council for Harmonization. Addendum on estimands and sensitivity analysis in clinical trials. 2019.

7. Yassi N, Hayward KS, Campbell BCV, Churilov L. Use of the estimand framework to manage the disruptive effects of covid-19 on stroke clinical trials. Stroke. 2021;52:3739–3747.

8. Kwakkel G, Lannin NA, Borschmann K, English C, Ali M, Churilov L, et al. Standardized measurement of sensorimotor recovery in stroke trials: Consensus-based core recommendations from the Stroke Recovery and Rehabilitation Roundtable. Neurorehabilitation and Neural Repair. 2017;31:784–792

9. Master Protocols for Drug and Biological Product Development Guidance for Industry U.S. Department of Health and Human Services Food and Drug Administration, Center for Drug Evaluation and Research (CDER) Center for Biologics Evaluation and Research (CBER) December 2023 Biostatistics / Clinical / Medical

10. White IR, Horton, N J. HN, Carpenter J, Pocock SJ. Strategy for intention to treat analysis in randomised trials with missing outcome data. British Medical Journal. 2011;342.

11. Viktorisson A, Buvarp D, Reinholdsson M, Danielsson A, Palstam A, Stibrant Sunnerhagen K. Associations of prestroke physical activity with stroke severity and mortality after intracerebral hemorrhage compared with ischemic stroke. Neurology, 2022;99(19):e2137–48.

12. Theou O, Pérez-Zepeda MU, van der Valk AM, Searle SD, Howlett SE, Rockwood K. A classification tree to assist with routine scoring of the Clinical Frailty Scale. Age and ageing. 2021 Jul;50(4):1406–11.

13. Yan, X., Lee, S., Li, N. Missing Data Handling Methods in Medical Device Clinical Trials Journal of Biopharmaceutical Statistics. 2009; 19:6, 1085–1098

14. Sui Y, Bu X, Duan Y, Li Y, Wang X. Application of Tipping Point Analysis in Clinical Trials using the Multiple Imputation Procedure in SAS. PharmaSUG 2023 – Paper SD-069.

15. USA Food and Drug Administration. Adaptive Designs for Clinical Trials of Drugs and Biologics: Guidance for Industry. 2019. https://www.fda.gov/media/78495/download

16. Marschner I.C. Confidence distributions for treatment effects in clinical trials: Posteriors without priors. Statistics in Medicine. 2024;43(6):1271–1289. doi: 10.1002/sim.10000

17. Johns H, Campbell B, Turc G, Churilov L. Power analysis for ordinal analyses of the modified Rankin scale and an online and downloadable tool for practical use. Stroke. 2023;54:1750–1760. doi: 10.1161/STROKEAHA.122.041260

18. Herdman M, Gudex C, Lloyd A, et al. Development and preliminary testing of the new five-level version of EQ-5D (EQ-5D-5L). Qual Life Res 2011;20:1727–36.

